# COVID19-related and all-cause mortality among middle-aged and older adults across the first epidemic wave of SARS-COV-2 infection in the region of Tarragona, Spain: results from the COVID19 TARRACO Cohort Study, March-June 2020

**DOI:** 10.1101/2021.02.02.21251028

**Authors:** Angel Vila-Corcoles, Eva Satue-Gracia, Angel Vila-Rovira, Cinta de Diego-Cabanes, Maria Jose Forcadell-Peris, Olga Ochoa-Gondar

## Abstract

**Background:** Direct and indirect COVID19-related mortality is uncertain. This study investigated COVID19-related and all-cause deaths among middle-aged and older adults during the first wave of COVID19 epidemic period, assessing mortality risks by pre-existing socio-demographic and medical underlying conditions.

**Methods:** Population-based cohort study involving 79,083 individuals ≥50 years-old in Tarragona (Southern Catalonia, Spain). Baseline cohort characteristics (age/sex, comorbidities and medications/vaccinations history) were established at study start (01/03/2020) and main outcomes were COVID19-related deaths (occurred in patients diagnosed with the disease) and all-cause deaths occurred among cohort members between 01/03/2020-30/06/2020. Mortality risks were assessed by Cox regression analyses.

**Results:** Cohort members were followed for 1,356,358 persons-weeks, occurring 576 all-cause deaths (124 COVID19-related). All-cause mortality rate was 42.5 deaths per 100,000 persons-week, being 22.8 in healthy/unrelated-COVID19 subjects, 236.4 in COVID19-excluded/PCR-negative subjects, 493.7 in COVID19-compatible/PCR-unperformed subjects and 4009.1 in COVID19-confirmed patients. In multivariable analyses, increasing age, sex male, nursing-home residence, cancer, neurologic, cardiac or liver disease, receiving diuretics, systemic corticosteroids, proton-pump inhibitors and benzodiazepines were associated with increased risk of all-cause mortality; conversely, receiving renin-angiotensin inhibitors and statins were associated with reduced risk. Age/years, sex male and nursing-home residence were strong predictors for COVID19-related mortality, but none comorbidity appeared significantly associated with an increased risk.

**Conclusion:** Apart from direct COVID19-related deaths (which represented almost 22% of all-cause mortality), theoretically COVID19-excluded patients (PCR-negative) suffered considerable greater all-cause mortality than healthy/unrelated-COVID19 subjects, which could explain, in part, the large excess deaths observed across the COVID19 pandemic.

## INTRODUCTION

A year ago, following an outbreak of pneumonia in Wuhan (China), a new coronavirus was detected. Said virus, later named Severe Acute Respiratory Syndrome Coronavirus 2 (SARS-CoV-2), is the cause of the disease designated by the World Health Organization (WHO) as COVID19 in February 2020.[1] Currently, the pandemic caused by this virus continues to be a threat to public health worldwide.[2]

Available publications have reported different percentages of asymptomatic or mild cases of COVID19 but, even if they account for around 80%,[2,3] due to the high infectivity of the virus, the number of disease’s related deaths is very high.[2] The mortality rates reported are very heterogeneous, depending on the countries (with important differences between regions, even within the same country), and also on the availability / policy of conducting diagnostic tests.[2]. For example, in Lombardy (Italy) the case fatality rate ranged between 1.6% and 18.3%,[4] while the Chinese Centre for Disease Control and Prevention, in a cohort of 72,314 cases, estimated a 2.3% case-fatality rate (14.8% for patients ≥80 years)[5]. Most articles report in-hospital mortality or case fatality rates (mortality rate among confirmed cases); it must be taken into account that infection fatality rate (which consider all infected individuals) would be lower. On the other hand, many deaths from COVID19 occur in undiagnosed patients. Lastly, we have to keep in mind the “excess mortality” (deaths caused by other conditions, linked to a delay in care, overburden of the health system and socioeconomic determinants of health).[6,7]

In the case of Spain, the latest scientific-technical report of the Ministry of Health (dated November 12) informed a fatality rate of 11% in patients ≥50 years (reaching 19% in the group ≥70 years compared to 0.45% in aged 50-69 years).[8] However, the estimates that took into account the results of the seroprevalence study (done in May)[9] reported much lower rates: 0.04% in the younger (50-69 years) and 4.1% in those ≥70 years, with a global rate for ≥50 years of 1.5%.[8] According to the Statistics National Institute (INE), as compared with 2019, almost a 20% increased all-cause death occurred in Spain during 2020 (surely as consequence of COVID19 pandemic).[10]

Most reviews agree that age and gender are well-established risk factors of mortality in COVID19 patients,[11,12] but there is no clear consensus on the specific contribution of pre-existing conditions to the risk of mortality from the disease.[13] Knowing the relative contribution of factors related to the patient (including socio-demographic and medical aspects) to mortality is very important to identify the individuals with greater risk and guarantee a more efficient use of resources.

This study investigated COVID19-related and all-cause mortality among middle-aged and older adults in the region of Tarragona across the first epidemic wave (March-June 2020), assessing the possible association between previous conditions (demographic, comorbidities, chronic medications’ use) and risk of death (COVID19-related and/or any cause) among middle-aged and older adults (who support the greatest burden of severe disease).

## METHODS

### Design, setting and study population

This is part of a large population-based retrospective-prospective cohort study (initiated in March 2020 and called *COVID19 TARRACO Cohort Study*) involving 79,083 individuals 50 years or older in the region of Tarragona (Southern Catalonia, Spain).

The design, setting and study population have been extensively described elsewhere, in prior articles that evaluated the epidemiology of COVID-19 during the first 8-12 weeks of epidemic period in the study area.[14,15]

The 79,083 cohort members were all individuals ≥50 years-old affiliated to the 12 Primary Care Centres (PCCs) managed by the *Institut Català de la Salut* in the study area (*Tarragonés, Alt Camp* and *Conca de Barberà* counties) and represented approximately 75% of overall inhabitants in this age strata according to census data.[16] This report focuses on cohort members who died (by COVID-19 or any cause) in the study setting between March 1 and June 30, 2020. The study was approved by the Ethical Committee of the Institution (Ethics Committee *IDIAP Jordi Gol*, Barcelona, file 20/065-PCV) and was conducted in accordance with the general principles for observational studies.[17] The study was determined to be exempt for informed consent under the public health surveillance exception.

### Data sources

Former CAPAMIS Research Database, an institutional clinical database used for earlier cohort studies in the region,[18] was updated and exploited as the prime data source in this COVID-19 epidemiological report. Summarily, this database collects inputs from the electronic system (e-CAP) of primary care clinical records, that includes administrative and clinical data, coded according to the International Classification of Diseases 10th Revision (ICD-10). It allowed us to identify socio-demographic aspects and previous conditions/comorbidities among cohort members in order to define their baseline profile at study onset (01/03/2020).

Two new electronic alerts, related to COVID-19’s laboratory results and ICD-10 codes for COVID-19 suspicion (B34.2, B97.29) were introduced to the e-CAP system at the beginning of the epidemic; later, both information sources were bonded to create the baseline research database utilised in this study.

Vital status was considered according to administrative data periodically updated in the e-CAP system. In addition, complementary data registered during emergency visits and/or hospital-stays, were used to complement vital status data in COVID-19 cases identified across study period. These data were transferred to a standardised form, after retrospective review of electronic records carried out by a research-trained group of family physicians and were afterwards linked with the prime research database containing baseline characteristics of 79,083 cohort members.

### Outcome definitions

Primary outcome was death from any cause (COVID19-related or not) occurred among cohort members during the 4-month of study period (01/03/2020-30/06/2020). Laboratory-confirmed COVID-19 case was defined when a cohort member tested positive in SARS-COV-2 using Reverse Transcription-Polymerase Chain Reaction (RT-PCR) or serological tests, according institutional guidelines.[19] COVID19-related death was considered when the patient died from any cause within the first 30-days after the onset of the disease or at any time during hospital-stay. Death by COVID-19 (likely due to COVID-19) or death with COVID-19 (likely due to other concomitant causes) were differentiated according to family physician’s criteria after checking hospital and primary care clinical records in each of deceased COVID19 cases.

### Covariates

Baseline covariates were age, sex, type of residence (community-dwelling or nursing-home), vaccinations’ history, previous comorbidities and chronic medications’ use; they were considered according to data registered in the e-CAP system.

Criteria used to define comorbidities and chronic medication use are extensively described in an appendix elsewhere.[15] Unrecorded comorbidities, medications or vaccinations were considered as absent.

In addition, at the end of the study period, according to their COVID-19 relation, cohort members were classified as: COVID19-unrelated/healthy, COVID19-excluded (PCR/serological test performed with negative result), COVID19-compatible (clinical suspicion alone, without any test performed) and COVID19-confirmed (positive PCR or serological test).

### Statistical analysis

Descriptive statistics of socio-demographic and clinical variables include frequencies/ percentages for categorical variables; and means/ standard deviations (SD) for continuous variables. Chi-squared or Fisher test (categorical variables) and Student’s t test (continuous variables) were used for comparison of differences between groups.

Mortality Rates (MRs) for COVID19-related and all-cause deaths were calculated per 100,000 persons-week, considering in the denominator the sum of the persons-time contributed for each cohort member during the study period (17.4 weeks). The confidence intervals (CIs) for the MRs were calculated assuming a Poisson distribution for uncommon events.

Cox regression analyses were used to calculate unadjusted, age&sex-adjusted and multivariable-adjusted hazards ratios (HRs) and estimate the association between baseline conditions and the time to the first outcome (COVID19-related or all-cause death) occurred among cohort members throughout the study period (from 1st March to 30th June 2020). All exposure above mentioned covariates (i.e., age, sex, residence, comorbidities/underlying conditions, chronic medications’ use and vaccinations’ history) were considered for multivariable Cox models; the method to select a subset of covariates to include in final models was the purposeful selection.[20] Final models include both significant or confounder variables and all covariates judged clinically or epidemiologically relevant. We performed a main analysis including the total study cohort (N=79,083) and two subgroup analyses restricted to community-dwelling individuals (N=77,669) and nursing-home residents (N=1414), which are shown in supplementary tables. The analyses were performed using IBM SPSS Statistics for Windows, version 24 (IBM Corp., Armonk, N.Y., USA); in all of them, statistical significance was set at p<0.05 (two-tailed).

## RESULTS

The study cohort included 79,083 persons (37,626 [47.6%] men and 41,407 [52.4%] women), with a mean age of 65.8 years (SD: 11.3). By age groups, 42,684 (54%) were aged 50-64 years and 10,386 (13.1%) were aged 80 years or older. A total of 77,669 (98.2%) were community-dwelling and 1414 (1.8%) nursing-home residents.

Cohort members were observed for a total of 1,356,358 persons-weeks (1,335,069 for community-dwelling and 21,289 for nursing-home residents). An amount of 4113 (5.2%) of the 79,083 cohort members was tested for SARS-COV-2 infection across study period; 3577 (87%) were negative and 536 (13%) positive (345 in community-dwelling and 191 in nursing-home residents). Two hundred and eighty-three cohort members (280 community-dwelling and 3 nursing-home residents) had a suspected COVID-19 without PCR testing (Table 1).

**Table 1.** Distribution of study population according to Covid19 related status at the end of study period. Population over 50 years, Tarragona region (Southern Catalonia, Spain), 01/03/2020-30/06/2020.

| COVID19 related status | COVID19<br>Nonrelated<br>(Healthy)<br>N=74687<br>n (%) | COVID19<br>Excluded<br>(PCR-)<br>N=3577<br>n (%) | COVID19<br>Suspected<br>(Without PCR)<br>N=283<br>n (%) | COVID19<br>Confirmed<br>(PCR+)<br>N=536<br>n (%) |  |
| --- | --- | --- | --- | --- | --- |
| Characteristic |  |  |  |  | p-value |
| Sociodemographical |  |  |  |  |  |
| Age: years, sd | 65.5 (11.1) | 70.0 (12.8) | 60.6 (9.4) | 74.0 (11.3) | <0.001 |
| Age group: 50-64 yrs | 40799 (54.6) | 1492 (41.7) | 216 (76.3) | 177 (33.0) | <0.001 |
| 65-79 yrs | 24638 (33.0) | 1186 (33.2) | 52 (18.4) | 137(25.6) |  |
| ≥80 yrs | 9250 (12.4) | 899 (25.1) | 15 (5.3) | 222 (41.4) |  |
| Sex Men | 35575 (47.6) | 1701 (47.6) | 115 (40.6) | 235 (43.8) | 0.036 |
| Women | 39112 (52.4) | 1876 (52.4) | 168 (59.4) | 301(56.2) |  |
| Community-dwelling | 73870 (98.9) | 3174 (88.7) | 280 (98.9) | 345 (64.4) | <0.001 |
| Nursing-home residence | 817 (1.1) | 403 (11.3) | 3 (1.1) | 191 (35.6) |  |
| Comorbidities |  |  |  |  |  |
| Neurological disease | 1935 (2.6) | 293 (8.2) | 10 (3.5) | 79 (14.7) | <0.001 |
| Renal disease | 3982 (5.3) | 420 (11.7) | 9 (3.2) | 65 (12.1) | <0.001 |
| Cancer | 5995 (8.0) | 549 (15.3) | 19 (6.7) | 67 (12.5) | <0.001 |
| Rheumatic disease | 804 (1.1) | 57 (1.6) | 4 (1.4) | 7 (1.3) | 0.032 |
| Inflammatory bowel disease | 497 (0.7) | 27 (0.8) | 2 (0.7) | 6 (1.1) | 0.567 |
| Respiratory disease | 6592 (8.8) | 567(15.9) | 33 (11.7) | 80 (14.9) | <0.001 |
| Cardiac disease | 12232 (16.4) | 1016 (28.4) | 34 (12.0) | 153 (28.5) | <0.001 |
| Atrial fibrillation | 3317 (4.4) | 392 (11.0) | 7 (2.5) | 70 (13.1) | <0.001 |
| Liver disease | 1338 (1.8) | 110 (3.1) | 6 (2.1) | 11 (2.1) | <0.001 |
| Diabetes | 12322 (16.5) | 838 (23.4) | 31 (11.0) | 126 (23.5) | <0.001 |
| Hypertension | 32604 (43.7) | 1937 (54.2) | 114 (40.3) | 290 (54.1) | <0.001 |
| Hypercholesterolemia | 25638 (34.3) | 1398 (39.1) | 97 (34.3) | 181 (33.8) | <0.001 |
| Obesity | 20468 (27.4) | 1013 (28.3) | 62 (21.9) | 135 (25.2) | 0.068 |
| Smoking | 12074 (16.2) | 591 (16.5) | 44 (15.5) | 41 (7.6) | <0.001 |
| Alcoholism | 1644 (2.2) | 134 (3.7) | 7 (2.5) | 11 (2.1) | <0.001 |
| Chronic medications use |  |  |  |  |  |
| Diuretics | 7586 (10.2) | 731 (20.4) | 25 (8.8) | 139 (25.9) | <0.001 |
| Beta blockers | 8817 (11.8) | 632 (17.7) | 35 (12.4) | 87 (16.2) | <0.001 |
| ACEIs | 15374 (20.6) | 872 (24.4) | 59 (20.8) | 114 (21.3) | <0.001 |
| ARBs | 8317 (11.1) | 471 (13.2) | 27 (9.5) | 54 (10.1) | 0.001 |
| Calcium channel blockers | 5990 (8.0) | 420 (11.7) | 16 (5.7) | 64 (11.9) | <0.001 |
| Statins | 15063 (20.2) | 936 (26.2) | 44 (15.5) | 91 (17.0) | <0.001 |
| Oral anticoagulants | 3461 (4.6) | 385 (10.8) | 8 (2.8) | 58 (10.8) | <0.001 |
| Antiplatelet drugs | 8344 (11.2) | 676 (18.9) | 26 (9.2) | 108 (20.1) | <0.001 |
| Insulin | 2726 (3.6) | 271 (7.6) | 2 (0.7) | 43 (8.0) | <0.001 |
| Oral antidiabetic drugs | 9855 (13.2) | 623 (17.4) | 22 (7.8) | 85 (15.9) | <0.001 |
| Inhaled respiratory drugs | 5628 (7.5) | 552 (15.4) | 36 (12.7) | 77 (14.4) | <0.001 |
| Antineoplastic agents | 104 (0.1) | 12 (0.3) | 0 (0.0) | 0 (0.0) | 0.017 |
| Systemic corticosteroids | 1105 (1.5) | 130 (3.6) | 7 (2.5) | 10 (1.9) | <0.001 |
| NSADs | 4079 (5.5) | 191 (5.3) | 30 (10.6) | 21 (3.9) | 0.001 |
| Chloroquine | 154 (0.2) | 11 (0.3) | 2 (0.7) | 1 (0.2) | 0.176 |
| Antihistamines | 3085 (4.1) | 152 (4.2) | 12 (4.2) | 15 (2.8) | 0.469 |
| Proton-Pump Inhibitors | 16254 (21.8) | 1423 (39.8) | 71 (25.1) | 183 (34.1) | <0.001 |
| Benzodiazepines | 12026 (16.1) | 819 (22.9) | 75 (26.5) | 126 (23.5) | <0.001 |
| Vaccination's history |  |  |  |  |  |
| Flu vaccine in prior autumn | 20838 (27.9) | 1449 (40.5) | 61 (21.6) | 258 (48.1) | <0.001 |
| PPV23 | 24189 (32.4) | 1666 (46.6) | 56 (19.8) | 272 (50.7) | <0.001 |
| PCV13 | 1005 (1.3) | 108 (3.0) | 8 (2.8) | 18 (3.4) | <0.001 |
| Tetanus | 47761 (63.9) | 2507 (70.1) | 187 (66.1) | 344 (64.2) | <0.001 |

NOTE: P-values were calculated by Chi-squared or Fisher's test as appropriate.

Across the four-month study period, 576 all-cause deaths were observed (413 in community-dwelling and 163 in nursing-home residents), of which 124 occurred among laboratory-confirmed COVID19 patients and were considered COVID19-related deaths (63 in community-dwelling and 61 in nursing-home residents). Of these 124 deceased patients with laboratory-confirmed COVID19, 112 (90.3%) died by (due to) COVID-19, while 12 (9.7%) died with COVID19 but likely due to other causes according to physician reviewers’ criteria after checking clinical records (ten cases in community dwelling and two cases in nursing-home residents).

All-cause mortality rate was 42.5 deaths per 100,000 persons-week (95% CI: 39.2-46.1) considering the total study cohort. By residence, it was 30.9 per 100,000 persons-week (95% CI: 28.0-34.1) in community-dwelling and 765.7 per 100,000 persons-week (95% CI: 653.9-896.6) in nursing-home residents.

COVID19-related mortality rate was 9.1 per 100,000 persons-week considering the total study cohort, being 4.7 per 100,000 persons-week among community-dwelling and 286.5 per 100,000 persons-week among nursing-home residents. Therefore, deaths from COVID19 represented a 21.5% (124/576) of the overall deaths observed in the total study cohort (15.3% [63/413] in community-dwelling vs 37.4% [61/163] in nursing-home residents).

Of the 576 all-cause deaths, 295 occurred among the 74,687 COVID19-unrelated/ healthy subjects (22.8 deaths per 100,000 persons-week), 150 among the 3577 COVID19-excluded/PCR-negative subjects (236.4 deaths per 100,000 persons-week), 7 among the 283 COVID19-suspected/PCR-unperformed (493.7 deaths per 100,000 persons-week) and 124 among the 536 COVID19-confirmed subjects (4009.1 deaths per 100,000 persons-week) (Table 2).

**Table 2.** Distribution of all-cause deaths according to COVID19-related status in cohort members. Tarragona, 01/03/2020-30/06/2020.

| **Mortality Population** | **No. of persons** | **Persons-time follow-up (No. of persons-week)** | **No. of deaths** | **Mortality Rate MR (95% CI)** |
| --- | --- | --- | --- | --- |
| **ALL PERSONS** | 79,083 | 1356358 | 576 | 42.5 (39.2-46.1) |
| Healthy (non-related COVID19) | 74,687 | 1290922 | 290 | 22.5 (20.1-25.2) |
| COVID19-excluded (PCR-) | 3577 | 60825 | 145 | 238.4 (201.2-282.3) |
| COVID19-suspected (non-PCR tested) | 283 | 1518 | 7 | 493.7 (198.0-1017.0) |
| COVID19-confirmed (PCR+) | 536 | 3093 | 124 | 4009.1 (3339.6-4810.9) |
| **COMMUNITY-DWELLING** | 77,669 | 1335069 | 413 | 30.9 (28.0-34.1) |
| Healthy (non-related COVID19) | 73,870 | 1277550 | 228 | 17.8 (15.7-20.2) |
| COVID19-excluded (PCR-) | 3174 | 54077 | 117-3 | 216.4 (180.3-259.7) |
| COVID19-suspected (non-PCR tested) | 280 | 1503 | 3-3 | 199.6 (41.1-582.8) |
| COVID19-confirmed (PCR+) | 345 | 1938 | 63 | 3250.8 (2503.1-4226.0) |
| **NURSING-HOME** | 1414 | 21289 | 163 | 765.7 (653.9-896.6) |
| Healthy (non-related COVID19) | 817 | 13371 | 67 | 501.1 (393.4-636.4) |
| COVID19-excluded (PCR-) | 403 | 6747 | 33-2 | 489.1 (340.9-679.8) |
| COVID19-suspected (non-PCR tested) | 3 | 15 | 2-2 | 13333.3 (1613.3-48133.2) |
| COVID19-confirmed (PCR+) | 191 | 1155 | 61 | 5281.4 (4066.7-6865.8) |

NOTE: MR denotes mortality rate, and it's calculated for 100,000 persons-week. CI denotes confidence interval.

Table 3 shows univariate analyses reporting all-cause deaths (n=576) by age, sex and pre-existing conditions (comorbidities, chronic medications’ use and vaccinations’ history) in the total study cohort. Table 4 reports it for COVID19-related deaths (n=124). Tables 5 and 6 show Cox regression analyses assessing unadjusted, age&sex-adjusted and multivariable-adjusted risks of suffer all-cause death and COVID19-related death, respectively, in the total study cohort.

**Table 3.** Incidence of all-cause mortality according to baseline demographical and clinical characteristics (comorbidities/medications) in the total study cohort (N=79,083). Tarragona region (Southern Catalonia, Spain), 01/03/2020-30/06/2020.

| Characteristic | Study population<br>(N=79083)<br>n (%) | All-cause deaths (n=576) |  |  |
| --- | --- | --- | --- | --- |
|  |  | Univariate analysis<br>n (%) p-value | Time follow-up<br>(persons-week) | Mortality rate |
|  |  |  |  | MR* (95% CI) |
| Sociodemographical |  |  |  |  |
| Age: 50-64 yrs | 42684 (54.0) | 54 (9.4) <0.001 | 734123 | 7.4 (5.5-9.8) |
| 65-79 yrs | 26013 (32.9) | 149 (25.9) | 447474 | 33.3 (28.1-39.4) |
| ≥80 yrs | 10386 (13.1) | 373 (64.8) | 174761 | 213.4 (191.8-237.3) |
| Sex Men | 37626 (47.6) | 302 (52.4) 0.019 | 645470 | 46.8 (41.7-52.5) |
| Women | 41457 (52.4) | 274 (47.6) | 710888 | 38.5 (34.0-43.7) |
| Community-dwelling | 77676 (98.2) | 413 (71.7) <0.001 | 1335069 | 30.9 (28.0-34.1) |
| Nursing-home residence | 1407 (1.8) | 163 (28.3) | 21289 | 765.7 (653.9-896.6) |
| Comorbidities |  |  |  |  |
| Neurological disease | 2317 (2.9) | 119 (20.7) <0.001 | 38344 | 310.3 (258.5-372.4) |
| Renal disease | 4476 (5.7) | 134 (23.3) <0.001 | 75629 | 177.2 (149.6-209.8) |
| Cancer | 6630 (8.4) | 168 (29.2) <0.001 | 112481 | 149.4 (127.6-174.9) |
| Rheumatic disease | 872 (1.1) | 10 (1.7) 0.144 | 14897 | 67.1 (32.2-123.5) |
| Inflammatory bowel disease | 532 (0.7) | 8 (1.4) 0.035 | 9097 | 87.9 (37.9-173.2) |
| Respiratory disease | 7272 (9.2) | 129 (22.4) <0.001 | 123794 | 104.2 (86.8-125.0) |
| Cardiac disease | 13435 (17.0) | 263 (45.7) <0.001 | 228716 | 115.0 (101.4-130.4) |
| Atrial fibrillation | 3786 (4.8) | 114 (19.8) <0.001 | 63962 | 178.2 (148.4-213.8) |
| Liver disease | 1465 (1.9) | 22 (3.8) <0.001 | 24998 | 88.0 (55.2-132.9) |
| Diabetes | 13317 (16.8) | 183 (31.8) <0.001 | 227640 | 80.4 (69.3-93.3) |
| Hypertension | 34945 (44.2) | 406 (70.5) <0.001 | 597890 | 67.9 (61.5-75.0) |
| Hypercholesterolemia | 27314 (34.5) | 228 (39.6) 0.011 | 468535 | 48.74 (43.0-55.2) |
| Obesity | 21678 (27.4) | 151 (26.2) 0.518 | 372086 | 40.6 (34.7-47.5) |
| Smoking | 12750 (16.1) | 60 (10.4) <0.001 | 219518 | 27.3 (21.0-35.5) |
| Alcoholism | 1796 (2.3) | 17 (3.0) 0.271 | 30774 | 55.2 (32.2-88.3) |
| Chronic medications use |  |  |  |  |
| Diuretics | 8481 (10.7) | 238 (41.3) <0.001 | 143357 | 166.0 (146.4-188.2) |
| Beta blockers | 9571 (12.1) | 138 (24.0) <0.001 | 163450 | 84.4 (71.2-99.9) |
| ACEIs | 16419 (20.8) | 142 (24.7) 0.021 | 281514 | 50.4 (42.5-59.7) |
| ARBs | 8869 (11.2) | 64 (11.1) 0.937 | 152376 | 42.0 (32.3-54.6) |
| Calcium channel blockers | 6490 (8.2) | 71 (12.3) <0.001 | 111107 | 63.9 (50.2-81.2) |
| Statins | 16134 (20.4) | 123 (21.4) 0.569 | 277237 | 44.4 (37.0-53.3) |
| Oral anticoagulants | 3912 (4.9) | 90 (15.6) <0.001 | 66424 | 135.5 (109.6-168.0) |
| Antiplatelet drugs | 9154 (11.6) | 173 (30.0) <0.001 | 155957 | 110.9 (95.6-128.6) |
| Insulin | 3042 (3.8) | 60 (10.4) <0.001 | 51741 | 116.0 (89.3-150.8) |
| Oral antidiabetic drugs | 10585 (13.4) | 123 (21.4) <0.001 | 181452 | 67.8 (56.5-81.4) |
| Inhaled respiratory drugs | 6293 (8.0) | 125 (21.7) <0.001 | 106857 | 117.0 (97.5-140.4) |
| Antineoplastic agents | 1614 (2.0) | 16 (2.8) 0.209 | 27691 | 57.8 (33.1-93.6) |
| Systemic corticosteroids | 1252 (1.6) | 56 (9.7) <0.001 | 21030 | 266.3 (205.1-346.2) |
| NSADs | 4321 (5.5) | 10 (1.7) <0.001 | 74152 | 13.5 (6.5-24.8) |
| Chloroquine | 168 (0.2) | 0 (0.0) 0.266 | 2867 | - |
| Antihistamines | 3264 (4.1) | 11 (1.9) 0.007 | 56243 | 19.6 (9.8-35.1) |
| Proton-Pump Inhibitors | 17931 (22.7) | 312 (54.2) <0.001 | 305566 | 102.1 (91.1-114.5) |
| Benzodiazepines | 13046 (16.5) | 166 (28.8) <0.001 | 222537 | 74.6 (63.7-87.4) |
| Vaccination's history |  |  |  |  |
| Flu vaccine in prior autumn | 22606 (28.6) | 355 (61.6) <0.001 | 385668 | 92.0 (82.7-102.3) |
| PPV23 | 26183 (33.1) | 395 (68.6) <0.001 | 447270 | 88.3 (80.0-97.5) |
| PCV13 | 1139 (1.4) | 24 (4.2) <0.001 | 19272 | 124.5 (79.8-185.5) |
| Tetanus | 50799 (64.2) | 415 (72.0) <0.001 | 871204 | 47.6 (43.1-52.6) |

NOTE: P-values in univariate analysis were calculated by chi-squared, or Fisher's test as appropriate, comparing percentages in the study population vs all-cause deaths cases; MR denotes mortality rates per 100.000 persons-week; CIs denotes confidence intervals for mortality rates and were calculated assuming a Poisson distribution for uncommon events.

**Table 4.** Incidence of Covid19-related mortality according to baseline demographical and clinical characteristics (comorbidities/medications) in the total study cohort (N=79,083). Tarragona region (Southern Catalonia, Spain), 01/03/2020-30/06/2020.

| Characteristic | Study population<br>(N=79083)<br>n (%) | COVID-19 deaths (n=124) |  |  |
| --- | --- | --- | --- | --- |
|  |  | Univariate analysis<br>n (%) p-value | Time follow-up<br>(persons-week) | Mortality rate |
|  |  |  |  | MR* (95% CI) |
| Sociodemographical |  |  |  |  |
| Age: 50-64 yrs | 42684 (54.0) | 3 (2.4) <0.001 | 734123 | 0.4 (0.1-1.2) |
| 65-79 yrs | 26013 (32.9) | 35 (28.2) | 447474 | 7.8 (5.4-10.8) |
| ≥80 yrs | 10386 (13.1) | 86 (69.4) | 174761 | 49.2 (39.8-61.0) |
| Sex Men | 37626 (47.6) | 63 (50.8) 0.471 | 645470 | 9.8 (7.5-12.7) |
| Women | 41457 (52.4) | 61 (49.2) | 710888 | 8.6 (6.6-11.2) |
| Community-dwelling | 77676 (98.2) | 63 (50.8) <0.001 | 1335069 | 4.7 (3.6-6.1) |
| Nursing-home residence | 1407 (1.8) | 61 (49.2) | 21289 | 286.5 (220.6-372.5) |
| Comorbidities |  |  |  |  |
| Neurological disease | 2317 (2.9) | 29 (23.4) <0.001 | 38344 | 75.6 (50.7-108.9) |
| Renal disease | 4476 (5.7) | 23 (18.5) <0.001 | 75629 | 30.4 (19.3-45.6) |
| Cancer | 6630 (8.4) | 23 (18.5) <0.001 | 112481 | 20.4 (12.9-30.6) |
| Rheumatic disease | 872 (1.1) | 0 (0.0) 0.239 | 14897 | - |
| Inflammatory bowel disease | 532 (0.7) | 2 (1.6) 0.200 | 9097 | 22.0 (2.7-79.4) |
| Respiratory disease | 7272 (9.2) | 27 (21.8) <0.001 | 123794 | 21.8 (14.4-31.8) |
| Cardiac disease | 13435 (17.0) | 54 (43.5) <0.001 | 228716 | 23.6 (17.5-31.2) |
| Atrial fibrillation | 3786 (4.8) | 23 (18.5) <0.001 | 63962 | 36.0 (22.8-54.0) |
| Liver disease | 1465 (1.9) | 3 (2.4) 0.639 | 24998 | 12.0 (2.5-35.0) |
| Diabetes | 13317 (16.8) | 41 (33.1) <0.001 | 227640 | 18.0 (12.9-24.5) |
| Hypertension | 34945 (44.2) | 79 (63.7) <0.001 | 597890 | 13.2 (10.5-16.5) |
| Hypercholesterolemia | 27314 (34.5) | 48 (38.7) 0.328 | 468535 | 10.2 (7.6-13.5) |
| Obesity | 21678 (27.4) | 30 (24.2) 0.421 | 372086 | 8.1 (5.5-11.6) |
| Smoking | 12750 (16.1) | 11 (8.9) 0.028 | 219518 | 5.0 (2.5-9.0) |
| Alcoholism | 1796 (2.3) | 3 (2.4) 0.912 | 30774 | 9.7 (2.0-28.3) |
| Chronic medications use |  |  |  |  |
| Diuretics | 8481 (10.7) | 49 (39.5) <0.001 | 143357 | 34.2 (25.4-45.1) |
| Beta blockers | 9571 (12.1) | 29 (23.4) <0.001 | 163450 | 17.7 (11.9-25.5) |
| ACEIs | 16419 (20.8) | 32 (25.8) 0.166 | 281514 | 11.4 (7.7-16.3) |
| ARBs | 8869 (11.2) | 11 (8.9) 0.408 | 152376 | 7.2 (3.6-12.9) |
| Calcium channel blockers | 6490 (8.2) | 18 (14.5) 0.010 | 111107 | 16.2 (9.6-25.6) |
| Statins | 16134 (20.4) | 26 (21.0) 0.876 | 277237 | 9.4 (6.1-13.8) |
| Oral anticoagulants | 3912 (4.9) | 17 (13.7) 0.001 | 66424 | 25.6 (14.9-41.0) |
| Antiplatelet drugs | 9154 (11.6) | 35 (28.2) <0.001 | 155957 | 22.4 (15.6-31.1) |
| Insulin | 3042 (3.8) | 12 (9.7) 0.001 | 51741 | 23.2 (12.0-40.6) |
| Oral antidiabetic drugs | 10585 (13.4) | 33 (26.6) <0.001 | 181452 | 18.2 (12.7-25.3) |
| Inhaled respiratory drugs | 6293 (8.0) | 28 (22.6) <0.001 | 106857 | 26.2 (17.4-38.0) |
| Antineoplastic agents | 1614 (2.0) | 4 (3.2) 0.350 | 27691 | 14.4 (3.9-36.9) |
| Systemic corticosteroids | 1252 (1.6) | 6 (4.8) 0.004 | 21030 | 28.5 (10.5-62.1) |
| NSADs | 4321 (5.5) | 0 (0.0) 0.007 | 74152 | - |
| Chloroquine | 168 (0.2) | 0 (0.0) 0.607 | 2867 | - |
| Antihistamines | 3264 (4.1) | 0 (0.0) 0.021 | 56243 | - |
| Proton-Pump Inhibitors | 17931 (22.7) | 56 (45.2) <0.001 | 305566 | 18.3 (14.1-23.8) |
| Benzodiazepines | 13046 (16.5) | 37 (29.8) <0.001 | 222537 | 16.6 (11.6-23.1) |
| Vaccination's history |  |  |  |  |
| Flu vaccine in prior autumn | 22606 (28.6) | 88 (71.0) <0.001 | 385668 | 22.8 (18.4-28.3) |
| PPV23 | 26183 (33.1) | 86 (69.4) <0.001 | 447270 | 19.2(15.5-23.8) |
| PCV13 | 1139 (1.4) | 4 (3.2) 0.095 | 19272 | 20.8 (5.7-53.2) |
| Tetanus | 50799 (64.2) | 82 (66.1) 0.660 | 871204 | 9.4 (7.5-11.8) |

NOTE: P-values in univariate analysis were calculated by chi-squared, or Fisher's test as appropriate, comparing percentages in the study population vs Covid19-related cases; MR denotes mortality rates per 100,000 persons-week; CIs denotes confidence intervals for mortality rates and were calculated assuming a Poisson distribution for uncommon events.

**Table 5.** Cox regression analyses assessing unadjusted, age & sex-adjusted and multivariable-adjusted risk of all-cause mortality in the total study cohort (N=79,083). Tarragona region (Southern Catalonia, Spain) from 01/03/2020 to 30/06/2020.

| Characteristic | All-cause deaths (n=576) |  |  |
| --- | --- | --- | --- |
|  | Unadjusted<br>HR (95% CI) p-value | Age & sex adjusted<br>HR (95% CI) p-value | Multivariable<br>HR (95% CI) p-value |
| **Sociodemographical** |  |  |  |
| **Age (continuous yrs)** | 1.12 (1.12-1.13) <0.001 | 1.13 (1.12-1.14) <0.001 | 1.08 (1.07-1.09) <0.001 |
| **Sex: women** | 0.82 (0.70-0.97) 0.020 | 0.58 (0.49-0.68) <0.001 | 0.68 (0.56-0.81) <0.001 |
| **Nursing-home residence** | 24.37 (20.33-29.22) <0.001 | 6.34 (5.18-7.76) <0.001 | 1.63 (1.28-2.09) <0.001 |
| **Comorbidities** |  |  |  |
| **Neurological disease** | 8.90 (7.28-10.89) <0.001 | 2.55 (2.06-3.15) <0.001 | 1.48 (1.18-1.85) 0.001 |
| **Renal disease** | 5.13 (4.22-6.22) <0.001 | 1.42 (1.17-1.74) 0.001 | 1.18 (0.95-1.46) 0.130 |
| **Cancer** | 4.55 (3.80-5.44) <0.001 | 2.64 (2.20-3.17) <0.001 | 2.46 (2.03-2.98) <0.001 |
| **Rheumatic disease** | 1.59 (0.85-2.97) 0.146 | 1.42 (0.76-2.65) 0.276 | 0.89 (0.46-1.71) 0.717 |
| **Respiratory disease** | 2.87 (2.36-3.49) <0.001 | 1.75 (1.43-2.13) <0.001 | 1.26 (0.96-1.66) 0.098 |
| **Cardiac disease** | 4.14 (3.51-4.88) <0.001 | 1.69 (1.43-2.00) <0.001 | 1.29 (1.06-1.56) 0.009 |
| **Atrial fibrillation** | 4.98 (4.06-6.11) <0.001 | 1.50 (1.22-1.85) <0.001 | 0.87 (0.63-1.19) 0.384 |
| **Liver disease** | 2.12 (1.38-3.24) 0.001 | 2.68 (1.75-4.10) <0.001 | 1.76 (1.14-2.73) 0.012 |
| **Diabetes** | 2.31 (1.94-2.75) <0.001 | 1.42 (1.19-1.69) <0.001 | 1.07 (0.80-1.44) 0.652 |
| **Hypertension** | 3.03 (2.53-3.62) <0.001 | 1.08 (0.90-1.30) 0.399 | 1.19 (0.95-1.48) 0.125 |
| **Obesity** | 0.94 (0.78-1.13) 0.513 | 0.92 (0.76-1.11) 0.274 | 0.89 (0.73-1.08) 0.219 |
| **Smoking** | 0.60 (0.46-0.79) <0.001 | 1.79 (1.35-2.38) <0.001 | 1.30 (0.97-1.74) 0.078 |
| **Alcoholism** | 1.31 (0.81-2.12) 0.273 | 2.14 (1.31-3.49) 0.002 | 1.42 (0.86-2.36) 0.173 |
| **Chronic medications use** |  |  |  |
| **Diuretics** | 5.95 (5.04-7.02) <0.001 | 2.22 (1.86-2.64) <0.001 | 1.55 (1.27-1.88) <0.001 |
| **Beta blockers** | 2.30 (1.90-2.78) <0.001 | 1.36 (1.12-1.64) 0.002 | 1.11 (0.90-1.37) 0.351 |
| **ACEIs** | 1.25 (1.03-1.51) 0.021 | 0.73 (0.60-0.88) 0.001 | 0.64 (0.52-0.79) <0.001 |
| **ARBs** | 0.99 (0.76-1.28) 0.927 | 0.59 (0.46-0.77) <0.001 | 0.53 (0.40-0.71) <0.001 |
| **Calcium channel blockers** | 1.58 (1.23-2.02) <0.001 | 0.83 (0.65-1.07) 0.151 | 0.81 (0.62-1.05) 0.114 |
| **Statins** | 1.06 (0.87-1.29) 0.582 | 0.72 (0.59-0.88) 0.002 | 0.73 (0.57-0.92) 0.008 |
| **Oral anticoagulants** | 3.59 (2.87-4.50) <0.001 | 1.27 (1.01-1.59) 0.041 | 1.10 (0.77-1.57) 0.594 |
| **Antiplatelet drugs** | 3.30 (2.76-3.95) <0.001 | 1.37 (1.14-1.64) 0.001 | 1.20 (0.97-1.49) 0.096 |
| **Insulin** | 2.93 (2.24-3.83) <0.001 | 1.88 (1.44-2.46) <0.001 | 1.15 (0.84-1.59) 0.376 |
| **Oral antidiabetic drugs** | 1.76 (1.44-2.15) <0.001 | 1.19 (0.97-1.45) 0.089 | 1.19 (0.87-1.62) 0.270 |
| **Inhaled respiratory drugs** | 3.24 (2.66-3.95) <0.001 | 1.77 (1.45-2.16) <0.001 | 1.08 (0.82-1.44) 0.575 |
| **Antineoplastic agents** | 1.37 (0.83-2.25) 0.213 | 1.40 (0.85-2.30) 0.187 | 0.63 (0.37-1.06) 0.082 |
| **Systemic corticosteroids** | 6.81 (5.17-8.98) <0.001 | 4.02 (3.05-5.30) <0.001 | 3.49 (2.59-4.70) <0.001 |
| **NSADs** | 0.31 (0.16-0.57) <0.001 | 0.52 (0.28-0.98) 0.043 | 0.65 (0.35-1.23) 0.189 |
| **Antihistamines** | 0.45 (0.25-0.82) <0.001 | 0.52 (0.29-0.95) 0.034 | 0.58 (0.32-1.05) 0.073 |
| **Proton-Pump Inhibitors** | 4.06 (3.45-4.78) <0.001 | 1.81 (1.53-2.14) <0.001 | 1.25 (1.03-1.51) 0.026 |
| **Benzodiazepines** | 2.06 (1.72-2.47) <0.001 | 1.45 (1.20-1.74) <0.001 | 1.34 (1.11-1.61) 0.003 |
| **Vaccination's history** |  |  |  |
| **Flu vaccine in prior autumn** | 4.04 (3.42-4.78) <0.001 | 1.24 (1.04-1.48) 0.017 | 1.03 (0.84-1.27) 0.761 |
| **PPV23** | 4.43 (3.72-5.29) <0.001 | 0.90 (0.75-1.09) 0.287 | 0.92 (0.72-1.18) 0.517 |
| **PCV13** | 3.01 (2.00-4.53) <0.001 | 1.81 (1.20-2.72) 0.005 | 1.13 (0.74-1.73) 0.577 |
| **Tetanus** | 1.44 (1.20-1.72) <0.001 | 0.83 (0.69-0.99) 0.042 | 0.88 (0.70-1.10) 0.263 |
| **Covid19-related status** |  |  |  |
| **Cas_4C: Covid19-nonrelated** | Reference (-) <0.001 | Reference (-) <0.001 | Reference (-) <0.001 |
| Covid19-negative | 10.78 (8.86-13.12) <0.001 | 6.44 (5.28-7.87) <0.001 | 4.24 (3.43-5.25) <0.001 |
| Covid19-suspected | 21.29 (10.01-45.30) <0.001 | 35.89 (16.86-76.42) <0.001 | 33.25 (15.53-71.20) <0.001 |
| Covid19-confirmed | 181.20 (144.29-227.54) <0.001 | 74.31 (58.49-94.40) <0.001 | 45.97 (35.01-60.37) <0.001 |

NOTE: HRs denotes Hazard ratios, and were calculated for those who had the condition as compared with those who had not the condition. In multivariable analysis the HRs were adjusted for age (continuous years), sex, residence, pre-existing comorbidities/underlying conditions, chronic medications use, vaccination's history and Covid19-related status across study period. CIs denote confidence intervals.

**Table 6.** Cox regression analyses assessing unadjusted, age & sex-adjusted and multivariable-adjusted risk of Covid19-related mortality in the total study cohort (N=79,083). Tarragona region (Southern Catalonia, Spain) from 01/03/2020 to 30/06/2020.

| Characteristic | COVID-19 deaths (n=124) |  |  |
| --- | --- | --- | --- |
|  | Unadjusted<br>HR (95% CI) p-value | Age & sex adjusted<br>HR (95% CI) p-value | Multivariable<br>HR (95% CI) p-value |
| **Sociodemographical** |  |  |  |
| **Age (continuous yrs)** | 1.13 (1.12-1.15) <0.001 | 1.14 (1.12-1.16) <0.001 | 1.08 (1.06-1.10) <0.001 |
| **Sex: women** | 0.88 (0.62-1.25) 0.472 | 0.60 (0.42-0.85) 0.004 | 0.55 (0.37-0.81) 0.003 |
| **Nursing-home residence** | 57.04 (40.11-81.11) <0.001 | 17.05 (11.28-25.78) <0.001 | 12.56 (8.07-19.54) <0.001 |
| **Comorbidities** |  |  |  |
| **Neurological disease** | 10.30 (6.79-15.61) <0.001 | 2.74 (1.77-4.23) <0.001 | 1.35 (0.86-2.14) 0.198 |
| **Renal disease** | 3.83 (2.44-6.03) <0.001 | 0.99 (0.62-1.57) 0.952 | 0.98 (0.60-1.60) 0.943 |
| **Cancer** | 2.51 (1.59-3.94) <0.001 | 1.43 (0.90-2.25) 0.130 | 1.39 (0.86-2.23) 0.178 |
| **Rheumatic disease** | 0.05 (0.01-92.92) 0.433 | 0.00 (-) 0.939 | 0.00 (-) 0.974 |
| **Respiratory disease** | 2.77 (1.81-4.24) <0.001 | 1.66 (1.08-2.55) 0.021 | 1.17 (0.66-2.08) 0.603 |
| **Cardiac disease** | 3.79 (2.66-5.41) <0.001 | 1.47 (1.02-2.11) 0.039 | 1.23 (0.81-1.85) 0.333 |
| **Atrial fibrillation** | 4.58 (2.91-7.20) <0.001 | 1.30 (0.82-2.07) 0.266 | 1.08 (0.54-2.16) 0.824 |
| **Liver disease** | 1.32 (0.42-4.15) 0.635 | 1.71 (0.54-5.37) 0.361 | 1.17 (0.37-3.77) 0.790 |
| **Diabetes** | 2.45 (1.68-3.56) <0.001 | 1.50 (1.03-2.19) 0.033 | 0.85 (0.41-1.77) 0.670 |
| **Hypertension** | 2.22 (1.54-3.21) <0.001 | 0.74 (0.51-1.07) 0.110 | 0.79 (0.50-1.24) 0.297 |
| **Obesity** | 0.84 (0.56-1.27) 0.420 | 0.84 (0.56-1.28) 0.418 | 0.89 (0.57-1.36) 0.580 |
| **Smoking** | 0.51 (0.27-0.94) 0.031 | 1.66 (0.87-3.18) 0.126 | 1.20 (0.61-2.34) 0.600 |
| **Alcoholism** | 1.07 (0.34-3.36) 0.911 | 1.92 (0.60-6.13) 0.272 | 1.07 (0.33-3.54) 0.910 |
| **Chronic medications use** |  |  |  |
| **Diuretics** | 5.49 (3.83-7.87) <0.001 | 1.88 (1.29-2.74) 0.001 | 1.48 (0.97-2.25) 0.068 |
| **Beta blockers** | 2.22 (1.47-3.37) <0.001 | 1.30 (0.86-1.97) 0.214 | 1.24 (0.78-1.98) 0.355 |
| **ACEIs** | 1.33 (0.89-1.99) 0.167 | 0.77 (0.51-1.15) 0.194 | 0.77 (0.49-1.22) 0.265 |
| **ARBs** | 0.77 (0.42-1.43) 0.408 | 0.46 (0.25-0.85) 0.013 | 0.52 (0.26-1.01) 0.054 |
| **Calcium channel blockers** | 1.90 (1.15-3.13) 0.012 | 0.99 (0.60-1.63) 0.970 | 1.14 (0.67-1.94) 0.633 |
| **Statins** | 1.04 (0.67-1.59) 0.877 | 0.73 (0.47-1.12) 0.147 | 0.91 (0.54-1.52) 0.709 |
| **Oral anticoagulants** | 3.08 (1.84-5.13) <0.001 | 1.05 (0.63-1.76) 0.854 | 0.87 (0.40-1.92) 0.734 |
| **Antiplatelet drugs** | 3.02 (2.04-4.46) <0.001 | 1.21 (0.81-1.80) 0.356 | 1.08 (0.67-1.74) 0.750 |
| **Insulin** | 2.69 (1.49-4.89) 0.001 | 1.72 (0.95-3.13) 0.073 | 1.01 (0.51-2.02) 0.970 |
| **Oral antidiabetic drugs** | 2.35 (1.58-3.50) <0.001 | 1.61 (1.08-2.40) 0.019 | 2.14 (1.03-4.43) 0.042 |
| **Inhaled respiratory drugs** | 3.40 (2.23-5.18) <0.001 | 1.81 (1.19-2.78) 0.006 | 1.45 (0.82-2.58) 0.207 |
| **Antineoplastic agents** | 1.60 (0.59-4.33) 0.355 | 1.65 (0.61-4.47) 0.325 | 1.55 (0.55-4.39) 0.412 |
| **Systemic corticosteroids** | 3.19 (1.41-7.25) 0.006 | 1.84 (0.81-4.19) 0.144 | 1.88 (0.80-4.41) 0.145 |
| **NSADs** | 0.05 (0.01-1.42) 0.078 | 0.00 (-) 0.949 | 0.00 (-) 0.968 |
| **Antihistamines** | 0.05 (0.01-2.38) 0.126 | 0.00 (-) 0.926 | 0.00 (-) 0.952 |
| **Proton-Pump Inhibitors** | 2.82 (1.98-4.12) <0.001 | 1.18 (0.83-1.69) 0.361 | 0.93 (0.61-1.41) 0.716 |
| **Benzodiazepines** | 2.16 (1.47-3.17) <0.001 | 1.48 (1.00-2.19) 0.052 | 1.40 (0.93-2.08) 0.104 |
| **Vaccination's history** |  |  |  |
| **Flu vaccine in prior autumn** | 6.14 (4.16-9.04) <0.001 | 1.82 (1.21-2.73) 0.004 | 1.55 (0.98-2.47) 0.063 |
| **PPV23** | 4.59 (3.13-6.72) <0.001 | 0.86 (0.58-1.29) 0.468 | 1.00 (0.59-1.67) 0.990 |
| **PCV13** | 2.30 (0.85-6.23) 0.101 | 1.37 (0.51-3.71) 0.537 | 1.15 (0.41-3.21) 0.794 |
| **Tetanus** | 1.09 (0.75-1.58) 0.659 | 0.62 (0.43-0.90) 0.013 | 0.68 (0.43-1.08) 0.103 |

NOTE: HRs denotes Hazard ratios, and were calculated for those who had the condition as compared with those who had not the condition. In multivariable analysis the HRs were adjusted for age (continuous years), sex, residence, pre-existing comorbidities/underlying conditions, chronic medications use and vaccination's history. CIs denote confidence intervals.

Supplementary tables S1-S4 show subgroup analyses restricted to community-dwelling individuals and supplementary tables S5-S8 show subgroup analyses restricted to nursing-home residents.

After multivariable-adjustments assessing risks of all-cause mortality in the total study cohort (see Table 5), increasing age/years (HR: 1.08; 95% CI: 1.07-1.09; p<0.001), male sex (HR: 1.47; 95% CI: 1.24-1.79; p<0.001), nursing-home residence (HR: 1.63; 95% CI: 1.28-2.09; p<0.001), neurological disease (HR: 1.48; 95% CI: 1.18-1.85; p=0.001), cancer (HR: 2.46; 95% CI: 2.03-2.98; p<0.001), cardiac disease (HR: 1.29; 95% CI: 1.06-1.56; p=0.009), liver disease (HR: 1.76; 95% CI: 1.14-2.73; p=0.012), receiving diuretics (HR: 1.55; 95% CI: 1.27-1.88; p<0.001), systemic corticosteroids (HR: 3.49; 95% CI: 2.59-4.70; p<0.001), proton-pump inhibitors (HR: 1.25; 95% CI: 1.03-1.51; p=0.026) and benzodiazepines (HR: 1.34; 95% CI: 1.11-1.61; p=0.003) were associated with a significant increased risk. Conversely, receiving Angiotensin Converting Enzyme inhibitors (ACEIs) (HR: 0.73; 95% CI: 0.60-0.88; p=0.001), Angiotensin II Receptor Blockers (ARBs) (HR: 0.59; 95% CI: 0.46-0.77; p<0.001) and statins (HR: 0.72; 95% CI: 0.59-0.88; p=0.002) were associated with a reduced risk. In subgroup analyses restricted to community-dwelling individuals, the results did not substantially change (see supplementary Table S3). In subgroup analysis restricted to nursing-home residents (see supplementary Table S7), history of tetanus vaccination was associated with a reduced risk (HR: 0.65; 95% CI: 0.44-0.97; p=0.036).

If we consider COVID19-related mortality (see Table 6), many comorbidities (cancer, neurological, renal, respiratory, cardiac disease, diabetes and hypertension) appeared associated with an increased risk in the crude analyses. However, after age&sex-adjustment only neurological, respiratory and cardiac disease were associated with increased risk, and none of them appeared significantly associated with an increased risk after multivariable-adjustment. In this analysis, increasing age/years (HR: 1.08; 95% CI: 1.06-1.10; p<0.001), male sex (HR: 1.82; 95% CI: 1.24-2.70; p=0.003) and nursing-home residence (HR: 12.56; 95% CI: 8.07-19.54; p<0.001) were the strongest predictors for COVID19-related mortality. Considering chronic medications’ use, while many medications were associated with an increased risk in crude analyses, none of them altered the risk of COVID19-related death in multivariable analysis (except for ARBs [HR: 0.52; 95% CI: 0.26-1.01; p=0.054] and oral antidiabetic drugs [HR: 2.14; 95% CI: 1.03-4.43; p=0.042], that were marginally associated with reduced and increased risk, respectively).

In subgroup analyses restricted to community-dwelling individuals evaluating risk of COVID19-related mortality, besides age/years and male sex, taking diuretics (HR: 1.89; 95% CI: 1.03-3.46; p=0.041) and systemic corticosteroids (HR: 3.10; 95% CI: 1.27-7.59; p=0.013) emerged associated with increased risk. None other medication, comorbidity and/or vaccination’s history altered the risk of COVID19-related death after multivariable-adjustments in these community-dwelling individuals (see supplementary Table S4).

In subgroup analyses restricted to nursing-home residents, apart from increasing age and male sex, only receiving antineoplastic agents (HR: 3.81; 95% CI: 1.00-14.44; p=0.049) and, surprisingly, influenza vaccination in prior autumn (HR: 2.34; 95% CI: 1.08-5.07; p=0.031) significantly altered the risk of COVID19-related death after multivariable adjustment (see supplementary Table S8).

## DISCUSSION

Direct and indirect mortality due to COVID19 is controversial.[4,5,8,11,13,21,22] This study investigated all-cause and COVID19-related deaths among middle-aged and older adults in the region of Tarragona (Southern Catalonia, Spain) across the first wave of COVID19 epidemic period (March-June 2020), assessing mortality risks by demographic characteristics and underlying medical conditions. Of note, the study was conducted in a region with relatively low incidence of COVID19 as compared with other European and Spanish regions.[2,3,8]

As main findings, all-cause mortality rate across study period was 42.5 deaths per 100,000 persons-week, and COVID19-related deaths represented 21.5% of overall deaths. Considering specifically COVID19-related deaths (those occurred among patients with laboratory-confirmed COVID19), 90.3% died by COVID-19 and 9.7% died with COVID19 (i.e., the death may be attributed to other baseline or concomitant cause.

Increasing age, male sex, and nursing-home residence appear as the strongest predictors for both COVID19-related death and all-cause death. Other pre-existing conditions may also alter mortality risk, especially some comorbidities and chronic medications’ use, which emerged significantly related with all-cause mortality risk.

Importantly, apart from COVID19-related deaths (occurring among patients with laboratory-confirmed COVID19), theoretically COVID19-excluded patients (i.e., those persons with a negative PCR) suffered ten times greater all-cause mortality than unrelated-COVID19/healthy subjects. This fact suggests that, apart from baseline excess risk in PCR-negative subjects (who were older and had more comorbidities than healthy/COVID19-unrelated subjects), some of the observed excess mortality in PCR-negative subjects could be due to undiagnosed/undetected COVID19 despite PCR performed. In this sense, we note that the reliability of PCR testing depends on the quality of the nasopharyngeal samples collected, timing of collection and sensitivity of tests used,[19] which could explain, in part, the excess all-cause mortality observed in PCR-negative subjects in this study.

Regarding excess mortality, the Statistics National Institute (INE) reported an increase in deceases in 2020 (compared to 2019) of 18.95%; which corresponds in absolute numbers to 70,715 deaths, of which only 43,131 were attributed to COVID19.[10] The considerably larger all-cause mortality in PCR-negative subjects observed in the present study may reflect COVID19 underdiagnosis and could explain, in part, some excess all-cause deaths observed across the current COVID19 pandemic.

We note the importance of providing adjusted data instead of crude data. Thus, while crude all-cause mortality rate was almost 25-times greater in nursing-home (766 deaths per 100,000 persons-week) than in community-dwelling individuals (31 deaths per 100,000 persons-week), we found that after multivariable adjustments (by age/sex and pre-existing conditions) nursing-home residence only increased approximately 1.6 times the adjusted risk of all-cause mortality (HR: 1.63; 95% CI: 1.28-2.09) as compared with community-dwelling. In this same way, apart from increasing age and sex male, many pre-existing conditions appeared significantly associated with an increased risk of COVID19-related mortality in the crude analysis (and several in the age&sex-adjusted analysis), but none comorbidity emerged significantly associated with an increased risk after multivariable adjustment.

This highlights the importance of maximizing adjusted data assessing event’s risk in observational studies. In fact, this could explain distinct/opposite data reported in different studies evaluating relationships between some pre-existing comorbidities/conditions and susceptibility/risk of suffering COVID19 infection or death.[4,5,11,13, 21-26] To illustrate it, besides subgroup analyses, we reported all unadjusted, age&sex-adjusted and multivariable-adjusted results here, which is a major strength in this study.

Considering chronic medication, there is scarce data reporting the possible influence of previous use of these drugs on the risk of COVID19-related mortality. We did not observe any significant association in this sense but, interestingly, receiving statins and renin-angiotensin inhibitors were significantly associated with a lower risk of all-cause mortality in our study cohort.

Major strengths in this study were the large size and representativity of the cohort (that included more than 79,000 people, which represents approximately 75% of overall population over 50 years of age in the study area, and the use of survival analysis methods to accurately estimate both risks of all-cause and COVID19-related mortality by distinct important population subgroups (based in its relation with COVID19): COVID19-confirmed subjects, COVID19-suspected subjects (without PCR performed), COVID19-excluded subjects (negative PCR testing) and COVID19-unrelated/healthy subjects). The large size of the study cohort together with the adjustment of major possible confounding variables (e.g., age, type of residence, pre-existing underlying conditions/comorbidities and chronic medications’ use) in the multivariable analyses may provide an acceptable basis to assess mortality risk among the general population ≥50 years across the first wave of COVID19 epidemic period in our setting.

Major limitations in this study are related with its retrospective design and scarce availability of PCR tests during the first weeks of study period. Indeed, considering that PCR testing was not routinely performed for all clinically compatible/suspected COVID19 patients across the study period, the laboratory-confirmed COVID19 cases (and, consequently, the number of COVID19-related deaths) were likely underestimated. Then, all-cause mortality (which is not influenced by the frequency of PCR testing) may be a better measure of COVID19 pandemic impact. As another limitation, the study was conducted in a single geographical area and, logically, specific mortality data may not be directly extrapolated to other geographical regions with distinct epidemic conditions.

The authors recognise these inherent limitations but note that, opposite to many papers reporting only crude COVID19 data, the present study provides age&sex-adjusted and multivariable-adjusted data evaluating both all-cause and COVID19-related mortality risks. Importantly, the estimations may considerably vary depending on type of analyses/adjustments performed; therefore, we underline again the importance of maximizing adjustments. We did subgroup analysis (nursing-home/community-dwelling) and multivariable-adjustments, but, as in all observational studies, a residual confounding due to unmeasured factors (e.g., socio-economical, lifestyle, job-or healthcare-related factors) cannot be excluded.

## CONCLUSIONS

In summary, in this large cohort study including 79,083 middle-aged and older adults followed across the first wave of COVID19 epidemic period in the region of Tarragona, we found that deaths from COVID19 represented 21.5% of all-cause mortality occurred across study period (March-June 2020). Increasing age, sex male, nursing-home residence, cancer, neurologic, cardiac or liver disease, receiving diuretics, systemic corticosteroids, proton-pump inhibitors and benzodiazepines were associated with an increased risk of all-cause mortality; conversely, receiving renin-angiotensin inhibitors and statins were associated with a reduced risk. Age/years, sex male and nursing-home residence were strong predictors for COVID19-related mortality, but no comorbidity was independently associated with increased risk.

Interestingly, apart from COVID19-related deaths (approximately 10% of them could be attributed to another baseline or concomitant cause), theoretically COVID19-excluded patients (PCR-negative) suffered considerable greater all-cause mortality than healthy/unrelated-COVID19 subjects. This data suggests COVID19 underdiagnosis and it could explain, in part, the unexpected excess deaths observed during the COVID19 pandemic.

## Data Availability

These data have been obtained from the Catalonian Health Institute Information System. Interested authors might obtain CAPAMIS data (previous ethics and scientific approval by the ethics and clinical research committee of the Primary Care Research Institute Jordi Gol (IDIAP Jordi Gol)) addressing purposes to the Institution.

## CONTRIBUTORS

AVC designed the study; AVC and ESG assessed outcomes and wrote the manuscript; ESG, CDC and MFP obtained data; ESG and AVR did statistical analyses and edited the manuscript; OOG revised the final version; AVC coordinated the study.

## SUPPLEMENTARY TABLES

**Supplementary Table S1.** Incidence of all-cause mortality according to baseline demographical and clinical characteristics (comorbidities/medications) in community-dwelling individuals (N=77,669). Tarragona region (Southern Catalonia, Spain), 01/03/2020-30/06/2020.

| Characteristic | Study population<br>(N=77669)<br>n (%) | All-cause deaths (n=413) |  |  |
| --- | --- | --- | --- | --- |
|  |  | Univariate analysis<br>n (%) p-value | Time follow-up<br>(persons-week) | Mortality rate |
|  |  |  |  | MR* (95% CI) |
| Sociodemographical |  |  |  |  |
| Age: 50-64 yrs | 42533 (54.8) | 52 (12.6) <0.001 | 731553 | 7.1 (5.3-9.4) |
| 65-79 yrs | 25712 (33.1) | 134 (32.4) | 442706 | 30.3 (25.6-35.9) |
| ≥80 yrs | 9424 (12.1) | 227 (55.0) | 160810 | 141.2 (124.5-160.1) |
| Sex Men | 37143 (47.8) | 237 (57.4) <0.001 | 638226 | 37.1 (32.7-42.1) |
| Women | 40526 (52.2) | 176 (42.6) | 696843 | 25.3 (21.8-29.3) |
| Comorbidities |  |  |  |  |
| Neurological disease | 1950 (2.5) | 61 (14.8) <0.001 | 33074 | 184.4 (142.0-239.7) |
| Renal disease | 4240 (5.5) | 98 (23.7) <0.001 | 72114 | 135.9 (111.2-165.8) |
| Cancer | 6461 (8.3) | 141 (34.1) <0.001 | 109968 | 128.2 (108.2-151.8) |
| Rheumatic disease | 860 (1.1) | 10 (2.4) 0.011 | 14699 | 68.0 (32.6-125.1) |
| Inflammatory bowel disease | 520 (0.7) | 7 (1.7) 0.010 | 8927 | 78.4 (31.4-161.5) |
| Respiratory disease | 7074 (9.1) | 104 (25.2) <0.001 | 120762 | 86.1 (70.4-105.0) |
| Cardiac disease | 12923 (16.6) | 190 (46.0) <0.001 | 221049 | 86.0 (74.1-99.8) |
| Atrial fibrillation | 3560 (4.6) | 80 (19.4) <0.001 | 60657 | 131.9 (105.3-164.9) |
| Liver disease | 1438 (1.9) | 21 (5.1) <0.001 | 24570 | 85.5 (52.9-130.8) |
| Diabetes | 12925 (16.6) | 130 (31.5) <0.001 | 221837 | 58.6 (48.8-70.3) |
| Hypertension | 33992 (43.8) | 291 (70.5) <0.001 | 583619 | 49.9 (44.5-55.9) |
| Hypercholesterolemia | 26765 (34.5) | 170 (41.2) 0.004 | 460087 | 36.9 (31.5-43.2) |
| Obesity | 21343 (27.5) | 122 (29.5) 0.347 | 366948 | 33.2 (27.7-39.8) |
| Smoking | 12640 (16.3) | 52 (12.6) 0.042 | 217755 | 23.9 (17.7-31.5) |
| Alcoholism | 1756 (2.3) | 15 (3.6) 0.060 | 30125 | 49.8 (27.9-82.2) |
| Chronic medications use |  |  |  |  |
| Diuretics | 8028 (10.3) | 161 (39.0) <0.001 | 136857 | 117.6 (100.4-137.7) |
| Beta blockers | 9311 (12.0) | 99 (24.0) <0.001 | 159529 | 62.1 (50.8-75.8) |
| ACEIs | 16030 (20.6) | 104 (25.2) 0.022 | 275658 | 37.7 (30.8-46.0) |
| ARBs | 8709 (11.2) | 52 (12.6) 0.374 | 149802 | 34.7 (25.7-45.8) |
| Calcium channel blockers | 6315 (8.1) | 56 (13.6) <0.001 | 108465 | 51.6 (39.7-67.1) |
| Statins | 15910 (20.5) | 107 (25.9) 0.006 | 273714 | 39.1 (32.0-47.7) |
| Oral anticoagulants | 3741 (4.8) | 66 (16.0) <0.001 | 63865 | 103.3 (81.1-131.2) |
| Antiplatelet drugs | 8809 (11.3) | 124 (30.0) <0.001 | 150896 | 82.2 (68.5-98.6) |
| Insulin | 2904 (3.7) | 41 (9.9) <0.001 | 49699 | 82.5 (58.9-112.2) |
| Oral antidiabetic drugs | 10352 (13.3) | 90 (21.8) <0.001 | 178039 | 50.6 (40.9-62.7) |
| Inhaled respiratory drugs | 6095 (7.8) | 103 (24.9) <0.001 | 103830 | 99.2 (81.1-121.0) |
| Antineoplastic agents | 1581 (2.0) | 11 (2.7) 0.365 | 27209 | 40.4 (20.2-72.3) |
| Systemic corticosteroids | 1216 (1.6) | 52 (12.6) <0.001 | 20457 | 254.2 (188.6-335.5) |
| NSADs | 4305 (5.5) | 9 (2.2) 0.003 | 73888 | 12.2 (5.6-23.2) |
| Antihistamines | 3221 (4.1) | 10 (2.4) 0.078 | 55545 | 18.0 (8.6-33.1) |
| Proton-Pump Inhibitors | 17315 (22.3) | 235 (56.9) <0.001 | 296238 | 79.3 (69.9-89.9) |
| Benzodiazepines | 12654 (16.3) | 120 (29.1) <0.001 | 216635 | 55.4 (46.1-66.5) |
| Vaccination's history |  |  |  |  |
| Flu vaccine in prior autumn | 21569 (27.8) | 223 (54.0) <0.001 | 370309 | 60.2 (52.3-69.3) |
| PPV23 | 25222 (32.5) | 282 (68.3) <0.001 | 432783 | 65.2 (58.2-73.1) |
| PCV13 | 1115 (1.4) | 21 (5.1) <0.001 | 18935 | 110.9 (68.6-169.7) |
| Tetanus | 49817 (64.1) | 318 (77.0) <0.001 | 856072 | 37.1 (33.1-41.6) |

NOTE: P-values in univariate analysis were calculated by chi-squared, or Fisher's test as appropriate, comparing percentages in the study population vs all-cause deaths cases; MR denotes mortality rates per 100.000 persons-week; CIs denotes confidence intervals for mortality rates and were calculated assuming a Poisson distribution for uncommon events.

**Supplementary Table S2.** Incidence of Covid19-related mortality according to baseline demographical and clinical characteristics (comorbidities/medications) in community-dwelling individuals (N=77,669). Tarragona region (Southern Catalonia, Spain), 01/03/2020-30/06/2020.

| Characteristic | Study population<br>(N=77669)<br>n (%) | COVID-19 deaths (n=63) |  |  |
| --- | --- | --- | --- | --- |
|  |  | Univariate analysis<br>n (%) p-value | Time follow-up<br>(persons-week) | Mortality rate |
|  |  |  |  | MR* (95% CI) |
| Sociodemographical |  |  |  |  |
| Age: 50-64 yrs | 42533 (54.8) | 2 (3.2) <0.001 | 731553 | 0.3 (0.1-1.1) |
| 65-79 yrs | 25712 (33.1) | 30 (47.6) | 442706 | 6.8 (4.6-9.7) |
| ≥80 yrs | 9424 (12.1) | 31 (49.2) | 160810 | 19.3 (13.0-27.6) |
| Sex Men | 37143 (47.8) | 35 (55.6) 0.219 | 638226 | 5.5 (3.8-7.6) |
| Women | 40526 (52.2) | 28 (44.4) | 696843 | 4.0 (2.7-5.8) |
| Comorbidities |  |  |  |  |
| Neurological disease | 1950 (2.5) | 8 (12.7) <0.001 | 33074 | 24.2 (10.4-47.7) |
| Renal disease | 4240 (5.5) | 13 (20.6) <0.001 | 72114 | 18.0 (9.6-30.8) |
| Cancer | 6461 (8.3) | 13 (20.6) <0.001 | 109968 | 11.8 (6.3-20.2) |
| Rheumatic disease | 860 (1.1) | 0 (0.0) 0.401 | 14699 | - |
| Inflammatory bowel disease | 520 (0.7) | 2 (3.2) 0.015 | 8927 | 22.4 (2.7-80.9) |
| Respiratory disease | 7074 (9.1) | 20 (31.7) <0.001 | 120762 | 16.6 (10.1-25.6) |
| Cardiac disease | 12923 (16.6) | 29 (46.0) <0.001 | 221049 | 13.1 (8.8-18.9) |
| Atrial fibrillation | 3560 (4.6) | 13 (20.6) <0.001 | 60657 | 21.4 (11.4-36.6) |
| Liver disease | 1438 (1.9) | 3 (4.8) 0.086 | 24570 | 12.2 (2.5-35.6) |
| Diabetes | 12925 (16.6) | 20 (31.7) 0.001 | 221837 | 9.0 (5.5-13.9) |
| Hypertension | 33992 (43.8) | 42 (66.7) <0.001 | 583619 | 7.2 (5.1-9.8) |
| Hypercholesterolemia | 26765 (34.5) | 24 (38.1) 0.544 | 460087 | 5.2 (3.3-7.7) |
| Obesity | 21343 (27.5) | 19 (30.2) 0.634 | 366948 | 5.2 (3.1-8.1) |
| Smoking | 12640 (16.3) | 5 (7.9) 0.073 | 217755 | 2.3 (0.7-5.4) |
| Alcoholism | 1756 (2.3) | 2 (3.2) 0.625 | 30125 | 6.6 (0.8-23.8) |
| Chronic medications use |  |  |  |  |
| Diuretics | 8028 (10.3) | 27 (42.9) <0.001 | 136857 | 19.7 (13.0-28.8) |
| Beta blockers | 9311 (12.0) | 18 (28.6) <0.001 | 159529 | 11.3 (6.7-17.9) |
| ACEIs | 16030 (20.6) | 15 (23.8) 0.534 | 275658 | 5.4 (3.0-8.9) |
| ARBs | 8709 (11.2) | 9 (14.3) 0.439 | 149802 | 6.0 (2.7-11.4) |
| Calcium channel blockers | 6315 (8.1) | 10 (15.9) 0.024 | 108465 | 9.2 (4.4-16.9) |
| Statins | 15910 (20.5) | 17 (27.0) 0.201 | 273714 | 6.2 (3.6-9.9) |
| Oral anticoagulants | 3741 (4.8) | 13 (20.6) <0.001 | 63865 | 20.4 (10.9-34.9) |
| Antiplatelet drugs | 8809 (11.3) | 16 (25.4) <0.001 | 150896 | 10.6 (6.1-17.2) |
| Insulin | 2904 (3.7) | 5 (7.9) 0.079 | 49699 | 10.1 (3.3-23.5) |
| Oral antidiabetic drugs | 10352 (13.3) | 17 (27.0) 0.001 | 178039 | 9.5 (5.5-15.2) |
| Inhaled respiratory drugs | 6095 (7.8) | 21 (33.3) <0.001 | 103830 | 20.2 (12.5-30.9) |
| Antineoplastic agents | 1581 (2.0) | 1 (1.6) 0.801 | 27209 | 3.7 (0.1-20.6) |
| Systemic corticosteroids | 1216 (1.6) | 6 (9.5) <0.001 | 20457 | 29.3 (10.8-63.9) |
| NSADs | 4305 (5.5) | 0 (0.0) 0.054 | 73888 | - |
| Antihistamines | 3221 (4.1) | 0 (0.0) 0.099 | 55545 | - |
| Proton-Pump Inhibitors | 17315 (22.3) | 33 (52.4) <0.001 | 296238 | 11.1(7.7-15.4) |
| Benzodiazepines | 12654 (16.3) | 20 (31.7) 0.001 | 216635 | 9.2 (5.6-14.2) |
| Vaccination's history |  |  |  |  |
| Flu vaccine in prior autumn | 21569 (27.8) | 35 (55.6) <0.001 | 370309 | 9.5 (6.6-13.2) |
| PPV23 | 25222 (32.5) | 45 (71.4) <0.001 | 432783 | 10.4 (7.6-13.9) |
| PCV13 | 1115 (1.4) | 3 (4.8) 0.026 | 18935 | 15.8 (3.3-46.1) |
| Tetanus | 49817 (64.1) | 46 (73.0) 0.142 | 856072 | 5.4 (3.9-7.2) |

NOTE: P-values in univariate analysis were calculated by chi-squared, or Fisher's test as appropriate, comparing percentages in the study population vs Covid19-related cases; MR denotes mortality rates per 100.000 persons-week; CIs denotes confidence intervals for mortality rates and were calculated assuming a Poisson distribution for uncommon events.

**Supplementary Table S3.** Cox regression analyses assessing unadjusted, age & sex-adjusted and multivariable-adjusted risk of all-cause mortality in community-dwelling individuals (N=77,669). Tarragona region (Southern Catalonia, Spain) from 01/03/2020 to 30/06/2020.

| Characteristic | All-cause deaths (n=413) |  |  |
| --- | --- | --- | --- |
|  | Unadjusted<br>HR (95% CI) p-value | Age & sex adjusted<br>HR (95% CI) p-value | Multivariable<br>HR (95% CI) p-value |
| **Sociodemographical** |  |  |  |
| Age (continuous yrs) | 1.11 (1.10-1.12) <0.001 | 1.11 (1.10-1.12) <0.001 | 1.09 (1.08-1.10) <0.001 |
| Sex: women | 0.68 (0.56-0.83) <0.001 | 0.52 (0.43-0.63) <0.001 | 0.63 (0.51-0.78) <0.001 |
| **Comorbidities** |  |  |  |
| Neurological disease | 6.81 (5.19-8.94) <0.001 | 2.25 (1.70-2.98) <0.001 | 1.67 (1.25-2.23) 0.001 |
| Renal disease | 5.44 (4.34-6.83) <0.001 | 1.66 (1.31-2.10) <0.001 | 1.25 (0.97-1.61) 0.085 |
| Cancer | 5.77 (4.71-7.07) <0.001 | 3.35 (2.72-4.12) <0.001 | 2.87 (2.32-3.55) <0.001 |
| Rheumatic disease | 2.23 (1.19-4.17) 0.012 | 2.00 (1.07-3.74) 0.031 | 1.12 (0.57-2.17) 0.747 |
| Respiratory disease | 3.38 (2.71-4.23) <0.001 | 2.09 (1.67-2.62) <0.001 | 1.23 (0.87-1.74) 0.246 |
| Cardiac disease | 4.29 (3.54-5.21) <0.001 | 1.89 (1.55-2.32) <0.001 | 1.35 (1.07-1.70) 0.011 |
| Atrial fibrillation | 5.05 (3.95-6.44) <0.001 | 1.63 (1.27-2.11) <0.001 | 1.31 (0.87-1.97) 0.202 |
| Liver disease | 2.86 (1.84-4.43) <0.001 | 3.56 (2.30-5.53) <0.001 | 2.43 (1.55-3.81) <0.001 |
| Diabetes | 2.31 (1.87-2.84) <0.001 | 1.41 (1.15-1.74) 0.001 | 1.22 (0.84-1.75) 0.296 |
| Hypertension | 3.07 (2.49-3.79) <0.001 | 1.21 (0.97-1.51) 0.087 | 1.29 (0.99-1.69) 0.063 |
| Obesity | 1.11 (0.90-1.37) 0.350 | 1.05 (0.85-1.29) 0.683 | 0.98 (0.78-1.22) 0.834 |
| Smoking | 0.74 (0.55-0.99) 0.042 | 1.83 (1.35-2.49) <0.001 | 1.59 (1.16-2.19) 0.004 |
| Alcoholism | 1.63 (0.98-2.73) 0.062 | 2.31 (1.37-3.90) 0.002 | 1.66 (0.97-2.86) 0.065 |
| **Chronic medications use** |  |  |  |
| Diuretics | 5.59 (4.59-6.81) <0.001 | 2.36 (1.91-2.91) <0.001 | 1.54 (1.21-1.95) <0.001 |
| Beta blockers | 2.32 (1.85-2.91) <0.001 | 1.39 (1.10-1.74) 0.005 | 0.99 (0.77-1.27) 0.947 |
| ACEIs | 1.29 (1.04-1.62) 0.023 | 0.76 (0.61-0.95) 0.015 | 0.59 (0.46-0.75) <0.001 |
| ARBs | 1.14 (0.85-1.52) 0.377 | 0.67 (0.50-0.90) 0.008 | 0.47 (0.34-0.65) <0.001 |
| Calcium channel blockers | 1.77 (1.34-2.35) <0.001 | 0.95 (0.71-1.26) 0.698 | 0.85 (0.63-1.14) 0.262 |
| Statins | 1.36 (1.09-1.69) 0.007 | 0.86 (0.69-1.07) 0.185 | 0.70 (0.54-0.91) 0.006 |
| Oral anticoagulants | 3.79 (2.91-4.93) <0.001 | 1.37 (1.05-1.80) 0.020 | 0.87 (0.55-1.37) 0.537 |
| Antiplatelet drugs | 3.37 (2.73-4.16) <0.001 | 1.45 (1.17-1.80) 0.001 | 1.14 (0.88-1.48) 0.322 |
| Insulin | 2.85 (2.07-3.94) <0.001 | 1.86 (1.35-2.57) <0.001 | 1.16 (0.79-1.69) 0.450 |
| Oral antidiabetic drugs | 1.81 (1.43-2.29) <0.001 | 1.18 (0.93-1.49) 0.166 | 0.98 (0.67-1.42) 0.895 |
| Inhaled respiratory drugs | 3.94 (3.15-4.92) <0.001 | 2.22 (1.77-2.78) <0.001 | 1.31 (0.92-1.86) 0.134 |
| Antineoplastic agents | 1.32 (0.72-2.40) 0.370 | 1.32 (0.72-2.40) 0.369 | 0.60 (0.32-1.12) 0.106 |
| Systemic corticosteroids | 9.24 (6.91-12.35) <0.001 | 5.59 (4.17-7.48) <0.001 | 3.60 (2.61-4.96) <0.001 |
| NSADs | 0.38 (0.20-0.74) 0.004 | 0.59 (0.30-1.14) 0.118 | 0.58 (0.30-1.13) 0.108 |
| Antihistamines | 0.57 (0.31-1.07) 0.081 | 0.64 (0.34-1.19) 0.159 | 0.52 (0.27-0.97) 0.040 |
| Proton-Pump Inhibitors | 4.63 (3.81-5.62) <0.001 | 2.25 (1.84-2.76) <0.001 | 1.60 (1.26-2.01) <0.001 |
| Benzodiazepines | 2.11 (1.71-2.61) <0.001 | 1.59 (1.28-1.98) <0.001 | 1.34 (1.07-1.67) 0.011 |
| **Vaccination's history** |  |  |  |
| Flu vaccine in prior autumn | 3.06 (2.52-3.71) <0.001 | 1.03 (0.84-1.27) 0.779 | 0.88 (0.70-1.13) 0.317 |
| PPV23 | 4.49 (3.65-5.52) <0.001 | 1.05 (0.83-1.32) 0.688 | 0.90 (0.67-1.21) 0.477 |
| PCV13 | 3.72 (2.40-5.77) <0.001 | 2.21 (1.42-3.43) <0.001 | 1.14 (0.72-1.82) 0.584 |
| Tetanus | 1.87 (1.49-2.36) <0.001 | 1.06 (0.84-1.34) 0.618 | 0.94 (0.72-1.23) 0.655 |

NOTE: HRs denotes Hazard ratios, and were calculated for those who had the condition as compared with those who had not the condition. In multivariable analysis the HRs were adjusted for age (continuous years), sex, pre-existing comorbidities/underlying conditions, chronic medications use and vaccination's history. CIs denote confidence intervals.

**Supplementary Table S4.** Cox regression analyses assessing unadjusted, age & sex-adjusted and multivariable-adjusted risk of Covid19-related mortality in community-dwelling individuals (N=77,669). Tarragona region (Southern Catalonia, Spain) from 01/03/2020 to 30/06/2020.

| Characteristic | COVID-19 deaths (n=63) |  |  |
| --- | --- | --- | --- |
|  | Unadjusted<br>HR (95% CI) p-value | Age & sex adjusted<br>HR (95% CI) p-value | Multivariable<br>HR (95% CI) p-value |
| **Sociodemographical** |  |  |  |
| **Age (continuous yrs)** | 1.11 (1.09-1.13) <0.001 | 1.11 (1.09-1.14) <0.001 | 1.08 (1.05-1.12) <0.001 |
| **Sex: women** | 0.73 (0.45-1.21) 0.221 | 0.56 (0.34-0.92) 0.023 | 0.63 (0.36-1.09) 0.099 |
| **Comorbidities** |  |  |  |
| **Neurological disease** | 5.69 (2.71-11.94) <0.001 | 1.86 (0.87-3.99) 0.111 | 1.45 (0.66-3.20) 0.354 |
| **Renal disease** | 4.53 (2.46-8.34) <0.001 | 1.36 (0.72-2.57) 0.343 | 1.04 (0.53-2.03) 0.914 |
| **Cancer** | 2.88 (1.57-5.31) 0.001 | 1.65 (0.89-3.07) 0.112 | 1.43 (0.76-2.68) 0.271 |
| **Rheumatic disease** | 0.05 (0.00-1914.01) 0.576 | 0.00 (-) 0.955 | 0.00 (-) 0.977 |
| **Respiratory disease** | 4.67 (2.75-7.93) <0.001 | 2.94 (1.72-5.04) <0.001 | 1.46 (0.63-3.39) 0.379 |
| **Cardiac disease** | 4.29 (2.61-7.04) <0.001 | 1.90 (1.14-3.19) 0.014 | 1.41 (0.78-2.56) 0.255 |
| **Atrial fibrillation** | 5.45 (2.96-10.03) <0.001 | 1.80 (0.95-3.38) 0.071 | 0.78 (0.25-2.45) 0.673 |
| **Liver disease** | 2.66 (0.84-8.49) 0.098 | 3.31 (1.04-10.59) 0.043 | 2.34 (0.71-7.69) 0.161 |
| **Diabetes** | 2.33 (1.37-3.97) 0.002 | 1.44 (0.84-2.45) 0.181 | 0.78 (0.25-2.43) 0.664 |
| **Hypertension** | 2.57 (1.52-4.45) <0.001 | 1.00 (0.58-1.73) 0.997 | 0.97 (0.50-1.91) 0.936 |
| **Obesity** | 1.14 (0.67-1.95) 0.635 | 1.07 (0.62-1.84) 0.808 | 0.93 (0.52-1.64) 0.796 |
| **Smoking** | 0.44 (0.18-1.10) 0.080 | 1.06 (0.41-2.72) 0.913 | 0.89 (0.34-2.35) 0.807 |
| **Alcoholism** | 1.42 (0.35-5.80) 0.626 | 2.05 (0.49-8.56) 0.325 | 1.68 (0.39-7.31) 0.488 |
| **Chronic medications use** |  |  |  |
| **Diuretics** | 6.54 (3.97-10.77) <0.001 | 2.80 (1.65-4.78) <0.001 | 1.89 (1.03-3.46) 0.041 |
| **Beta blockers** | 2.94 (1.70-5.08) <0.001 | 1.76 (1.02-3.05) 0.043 | 1.35 (0.73-2.49) 0.341 |
| **ACEIs** | 1.20 (0.67-2.15) 0.535 | 0.71 (0.40-1.27) 0.246 | 0.59 (0.30-1.14) 0.114 |
| **ARBs** | 1.32 (0.65-2.67) 0.441 | 0.78 (0.39-1.58) 0.492 | 0.53 (0.24-1.17) 0.114 |
| **Calcium channel blockers** | 2.13 (1.08-4.19) 0.028 | 1.14 (0.58-2.26) 0.699 | 1.12 (0.55-2.29) 0.753 |
| **Statins** | 1.44 (0.82-2.50) 0.203 | 0.92 (0.53-1.60) 0.766 | 0.74 (0.38-1.42) 0.364 |
| **Oral anticoagulants** | 5.17 (2.81-9.51) <0.001 | 1.93 (1.03-3.60) 0.040 | 1.38 (0.42-4.46) 0.595 |
| **Antiplatelet drugs** | 2.67 (1.51-4.70) 0.001 | 1.15 (0.64-2.05) 0.646 | 0.90 (0.45-1.81) 0.776 |
| **Insulin** | 2.23 (0.89-5.56) 0.086 | 1.46 (0.59-3.64) 0.418 | 0.92 (0.33-2.59) 0.879 |
| **Oral antidiabetic drugs** | 2.40 (1.38-4.19) 0.002 | 1.58 (0.90-2.76) 0.109 | 2.00 (0.64-6.21) 0.233 |
| **Inhaled respiratory drugs** | 5.91 (3.50-9.98) <0.001 | 3.41 (2.00-5.82) <0.001 | 2.03 (0.87-4.73) 0.102 |
| **Antineoplastic agents** | 0.78 (0.11-5.60) 0.801 | 0.77 (0.11-5.57) 0.798 | 0.58 (0.08-4.37) 0.592 |
| **Systemic corticosteroids** | 6.69 (2.89-15.52) <0.001 | 4.05 (1.74-9.42) 0.001 | 3.10 (1.27-7.59) 0.013 |
| **NSADs** | 0.046 (0.00-5.48) 0.206 | 0.00 (-) 0.961 | 0.00 (-) 0.969 |
| **Antihistamines** | 0.047 (0.00-11.44) 0.275 | 0.00 (-) 0.964 | 0.00 (-) 0.968 |
| **Proton-Pump Inhibitors** | 3.85 (2.35-6.31) <0.001 | 1.85 (1.11-3.08) 0.019 | 1.29 (0.71-2.32) 0.404 |
| **Benzodiazepines** | 2.40 (1.41-4.07) 0.001 | 1.79 (1.04-3.08) 0.037 | 1.60 (0.92-2.79) 0.098 |
| **Vaccination's history** |  |  |  |
| **Flu vaccine in prior autumn** | 3.26 (1.98-5.35) <0.001 | 1.12 (0.66-1.89) 0.686 | 0.90 (0.49-1.64) 0.721 |
| **PPV23** | 5.21 (3.01-8.99) <0.001 | 1.28 (0.69-2.36) 0.435 | 1.37 (0.62-3.04) 0.443 |
| **PCV13** | 3.46 (1.09-11.03) 0.036 | 2.07 (0.65-6.61) 0.221 | 1.11 (0.33-3.76) 0.862 |
| **Tetanus** | 1.51 (0.87-2.64) 0.144 | 0.86 (0.49-1.51) 0.595 | 0.63 (0.32-1.25) 0.186 |

NOTE: HRs denotes Hazard ratios, and were calculated for those who had the condition as compared with those who had not the condition. In multivariable analysis the HRs were adjusted for age (continuous years), sex, pre-existing comorbidities/underlying conditions, chronic medications use and vaccination's history. CIs denote confidence intervals.

**Supplementary Table S5.** Incidence of all-cause mortality according to baseline demographical and clinical characteristics (comorbidities/medications) in nursing-home residents (N=1,414). Tarragona region (Southern Catalonia, Spain), 01/03/2020-30/06/2020.

| Characteristic | Study population<br>(N=1414)<br>n (%) | All-cause deaths (n=163) |  |  |
| --- | --- | --- | --- | --- |
|  |  | Univariate analysis<br>n (%) p value | Time follow-up<br>(persons-week) | Mortality rate |
|  |  |  |  | MR* (95% CI) |
| Sociodemographical |  |  |  |  |
| Age: 50-64 yrs | 151 (10.7) | 2 (1.2) <0.001 | 2570 | 77.8 (9.4-280.9) |
| 65-79 yrs | 301 (21.3) | 15 (9.2) | 4768 | 314.6 (176.2-519.1) |
| ≥80 yrs | 962 (68.0) | 146 (89.6) | 13951 | 1046.5 (883.1-1239.1) |
| Sex: Men | 483 (34.2) | 65 (39.9) 0.102 | 7245 | 897.2 (704.3-1139.4) |
| Women | 931 (65.8) | 98 (60.1) | 14045 | 697.8 (570.8-851.3) |
| Comorbidities |  |  |  |  |
| Neurological disease | 367 (26.0) | 58 (35.6) 0.003 | 5270 | 1100.6 (847.5-1430.8) |
| Renal disease | 236 (16.7) | 36 (22.1) 0.050 | 3515 | 1024.2 (713.9-1423.6) |
| Cancer | 169 (12.0) | 27 (16.6) 0.054 | 2514 | 1074.0 (707.8-1568.0) |
| Rheumatic disease | 12 (0.8) | 0 (0.0) 0.209 | 199 | - |
| Inflammatory bowel disease | 12 (0.8) | 1 (0.6) 0.728 | 170 | 588.2 (14.9-3276.3) |
| Respiratory disease | 198 (14.0) | 25 (15.3) 0.602 | 3032 | 824.5 (533.5-1220.3) |
| Cardiac disease | 512 (36.2) | 73 (44.8) 0.015 | 7666 | 952.3 (747.6-1209.4) |
| Atrial fibrillation | 226 (16.0) | 34 (20.9) 0.071 | 3305 | 1028.7 (717.0-1429.9) |
| Liver disease | 27 (1.9) | 1 (0.6) 0.199 | 428 | 233.6 (5.9-1301.2) |
| Diabetes | 392 (27.7) | 53 (32.5) 0.146 | 5803 | 913.3 (677.7-1205.6) |
| Hypertension | 953 (67.4) | 115 (70.6) 0.361 | 14270 | 805.9 (671.3-967.1) |
| Hypercholesterolemia | 549 (38.8) | 58 (35.6) 0.366 | 8449 | 686.5 (528.6-892.5) |
| Obesity | 335 (23.7) | 29 (17.8) 0.060 | 5138 | 564.4 (378.1-812.7) |
| Smoking | 110 (7.8) | 8 (4.9) 0.146 | 1763 | 453.8 (195.6-894.0) |
| Alcoholism | 40 (2.8) | 2 (1.2) 0.190 | 649 | 308.2 (37.3-1112.6) |
| Chronic medications use |  |  |  |  |
| Diuretics | 453 (32.0) | 77 (47.2) <0.001 | 6501 | 1184.4 (945.2-1480.5) |
| Beta blockers | 260 (18.4) | 39 (23.9) 0.052 | 3921 | 994.6 (710.1-1352.7) |
| ACEIs | 389 (27.5) | 38 (23.3) 0.202 | 5856 | 648.9 (452.3-902.0) |
| ARBs | 160 (11.3) | 12 (7.4) 0.090 | 2574 | 466.2 (241.0-815.9) |
| Calcium channel blockers | 175 (12.4) | 15 (9.2) 0.191 | 2642 | 567.8 (318.0-936.9) |
| Statins | 224 (15.8) | 16 (9.8) 0.025 | 3523 | 454.2 (259.8-735.8) |
| Oral anticoagulants | 171 (12.1) | 24 (14.7) 0.273 | 2559 | 937.9 (601.2-1397.5) |
| Antiplatelet drugs | 345 (24.4) | 49 (30.1) 0.074 | 5061 | 968.2 (718.4-1278.0) |
| Insulin | 138 (9.8) | 19 (11.7) 0.386 | 2042 | 930.5 (560.2-1451.6) |
| Oral antidiabetic drugs | 233 (16.5) | 33 (20.2) 0.168 | 3413 | 966.9 (673.9-1344.0) |
| Inhaled respiratory drugs | 198 (14.0) | 22 (13.5) 0.843 | 3027 | 726.8 (455.7-1097.5) |
| Antineoplastic agents | 33 (2.3) | 5 (3.1) 0.509 | 482 | 1037.3 (336.1-2416.9) |
| Systemic corticosteroids | 36 (2.5) | 4 (2.5) 0.937 | 573 | 698.1 (189.9-1787.1) |
| NSADs | 16 (1.1) | 1 (0.6) 0.506 | 264 | 378.8 (9.6-2109.9) |
| Antihistamines | 43 (3.0) | 1 (0.6) 0.055 | 698 | 143.3 (3.6-798.2) |
| Proton-Pump Inhibitors | 616 (43.6) | 77 (47.2) 0.314 | 9327 | 825.6 (658.8-1032.0) |
| Benzodiazepines | 392 (27.7) | 46 (28.2) 0.880 | 5902 | 779.4 (568.2-1044.4) |
| Vaccination's history |  |  |  |  |
| Flu vaccine in prior autumn | 1037 (73.3) | 132 (81.0) 0.019 | 15359 | 859.4 (725.3-1017.5) |
| PPV23 | 961 (68.0) | 113 (69.3) 0.692 | 14487 | 780.0 (649.7-936.0) |
| PCV13 | 24 (1.7) | 3 (1.8) 0.880 | 337 | 890.2 (183.4-2599.4) |
| Tetanus | 982 (69.4) | 97 (59.5) 0.003 | 15132 | 641.0 (524.3-782.0) |

NOTE: P-values in univariate analysis were calculated by chi-squared, or Fisher's test as appropriate, comparing percentages in the study population vs all-cause deaths cases; MR denotes mortality rates per 100.000 persons-week; CIs denotes confidence intervals for mortality rates and were calculated assuming a Poisson distribution for uncommon events.

**Supplementary Table S6.**
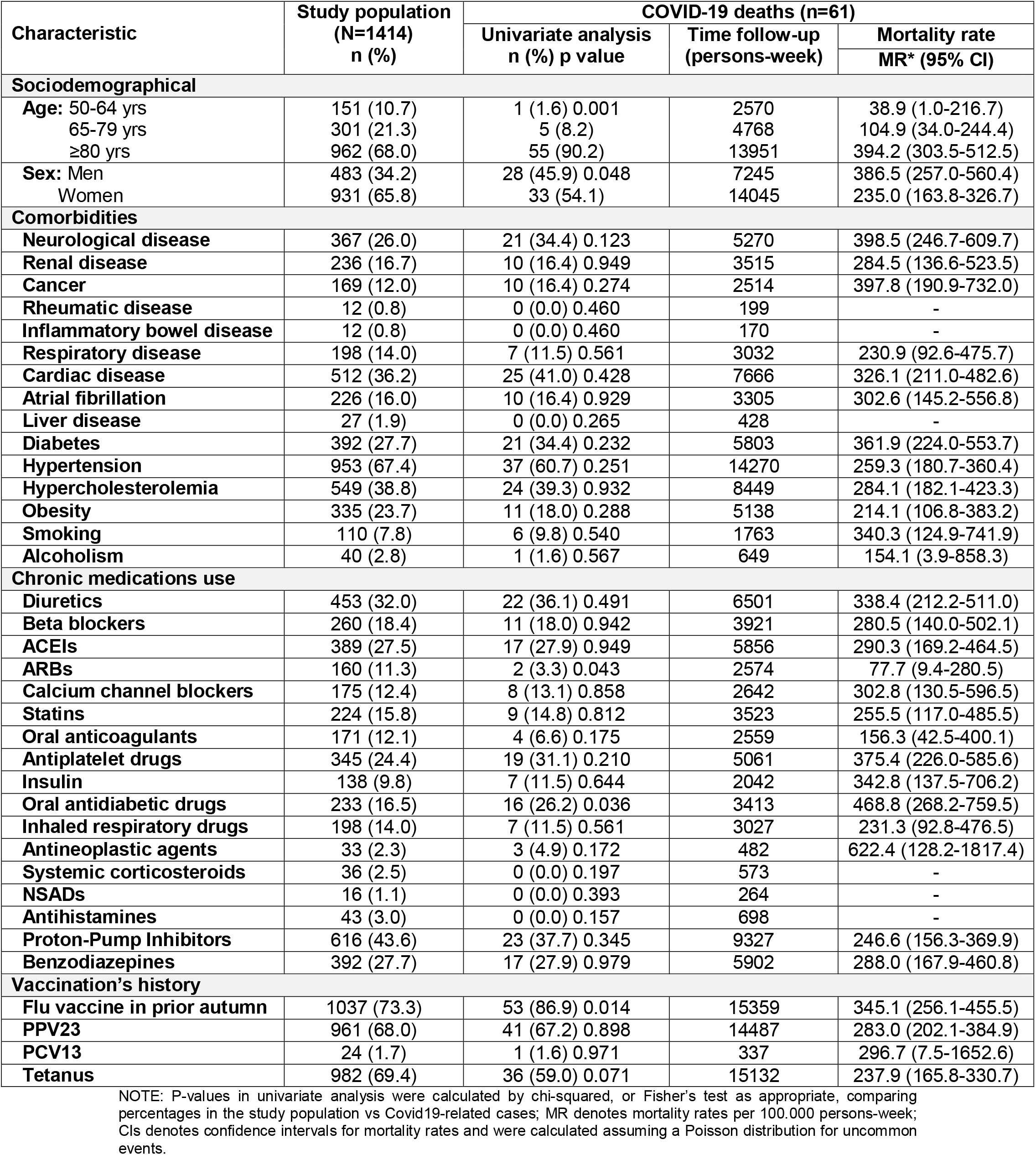
Incidence of Covid19-related mortality according to baseline demographical and clinical characteristics (comorbidities/medications) in nursing-home residents (N=1,414). Tarragona region (Southern Catalonia, Spain), 01/03/2020-30/06/2020.

| Characteristic | Study population<br>(N=1414)<br>n (%) | COVID-19 deaths (n=61) |  |  |
| --- | --- | --- | --- | --- |
|  |  | Univariate analysis<br>n (%) p value | Time follow-up<br>(persons-week) | Mortality rate |
|  |  |  |  | MR* (95% CI) |
| Sociodemographical |  |  |  |  |
| Age: 50-64 yrs | 151 (10.7) | 1 (1.6) 0.001 | 2570 | 38.9 (1.0-216.7) |
| 65-79 yrs | 301 (21.3) | 5 (8.2) | 4768 | 104.9 (34.0-244.4) |
| ≥80 yrs | 962 (68.0) | 55 (90.2) | 13951 | 394.2 (303.5-512.5) |
| Sex: Men | 483 (34.2) | 28 (45.9) 0.048 | 7245 | 386.5 (257.0-560.4) |
| Women | 931 (65.8) | 33 (54.1) | 14045 | 235.0 (163.8-326.7) |
| Comorbidities |  |  |  |  |
| Neurological disease | 367 (26.0) | 21 (34.4) 0.123 | 5270 | 398.5 (246.7-609.7) |
| Renal disease | 236 (16.7) | 10 (16.4) 0.949 | 3515 | 284.5 (136.6-523.5) |
| Cancer | 169 (12.0) | 10 (16.4) 0.274 | 2514 | 397.8 (190.9-732.0) |
| Rheumatic disease | 12 (0.8) | 0 (0.0) 0.460 | 199 | - |
| Inflammatory bowel disease | 12 (0.8) | 0 (0.0) 0.460 | 170 | - |
| Respiratory disease | 198 (14.0) | 7 (11.5) 0.561 | 3032 | 230.9 (92.6-475.7) |
| Cardiac disease | 512 (36.2) | 25 (41.0) 0.428 | 7666 | 326.1 (211.0-482.6) |
| Atrial fibrillation | 226 (16.0) | 10 (16.4) 0.929 | 3305 | 302.6 (145.2-556.8) |
| Liver disease | 27 (1.9) | 0 (0.0) 0.265 | 428 | - |
| Diabetes | 392 (27.7) | 21 (34.4) 0.232 | 5803 | 361.9 (224.0-553.7) |
| Hypertension | 953 (67.4) | 37 (60.7) 0.251 | 14270 | 259.3 (180.7-360.4) |
| Hypercholesterolemia | 549 (38.8) | 24 (39.3) 0.932 | 8449 | 284.1 (182.1-423.3) |
| Obesity | 335 (23.7) | 11 (18.0) 0.288 | 5138 | 214.1 (106.8-383.2) |
| Smoking | 110 (7.8) | 6 (9.8) 0.540 | 1763 | 340.3 (124.9-741.9) |
| Alcoholism | 40 (2.8) | 1 (1.6) 0.567 | 649 | 154.1 (3.9-858.3) |
| Chronic medications use |  |  |  |  |
| Diuretics | 453 (32.0) | 22 (36.1) 0.491 | 6501 | 338.4 (212.2-511.0) |
| Beta blockers | 260 (18.4) | 11 (18.0) 0.942 | 3921 | 280.5 (140.0-502.1) |
| ACEIs | 389 (27.5) | 17 (27.9) 0.949 | 5856 | 290.3 (169.2-464.5) |
| ARBs | 160 (11.3) | 2 (3.3) 0.043 | 2574 | 77.7 (9.4-280.5) |
| Calcium channel blockers | 175 (12.4) | 8 (13.1) 0.858 | 2642 | 302.8 (130.5-596.5) |
| Statins | 224 (15.8) | 9 (14.8) 0.812 | 3523 | 255.5 (117.0-485.5) |
| Oral anticoagulants | 171 (12.1) | 4 (6.6) 0.175 | 2559 | 156.3 (42.5-400.1) |
| Antiplatelet drugs | 345 (24.4) | 19 (31.1) 0.210 | 5061 | 375.4 (226.0-585.6) |
| Insulin | 138 (9.8) | 7 (11.5) 0.644 | 2042 | 342.8 (137.5-706.2) |
| Oral antidiabetic drugs | 233 (16.5) | 16 (26.2) 0.036 | 3413 | 468.8 (268.2-759.5) |
| Inhaled respiratory drugs | 198 (14.0) | 7 (11.5) 0.561 | 3027 | 231.3 (92.8-476.5) |
| Antineoplastic agents | 33 (2.3) | 3 (4.9) 0.172 | 482 | 622.4 (128.2-1817.4) |
| Systemic corticosteroids | 36 (2.5) | 0 (0.0) 0.197 | 573 | - |
| NSADs | 16 (1.1) | 0 (0.0) 0.393 | 264 | - |
| Antihistamines | 43 (3.0) | 0 (0.0) 0.157 | 698 | - |
| Proton-Pump Inhibitors | 616 (43.6) | 23 (37.7) 0.345 | 9327 | 246.6 (156.3-369.9) |
| Benzodiazepines | 392 (27.7) | 17 (27.9) 0.979 | 5902 | 288.0 (167.9-460.8) |
| Vaccination's history |  |  |  |  |
| Flu vaccine in prior autumn | 1037 (73.3) | 53 (86.9) 0.014 | 15359 | 345.1 (256.1-455.5) |
| PPV23 | 961 (68.0) | 41 (67.2) 0.898 | 14487 | 283.0 (202.1-384.9) |
| PCV13 | 24 (1.7) | 1 (1.6) 0.971 | 337 | 296.7 (7.5-1652.6) |
| Tetanus | 982 (69.4) | 36 (59.0) 0.071 | 15132 | 237.9 (165.8-330.7) |

NOTE: P-values in univariate analysis were calculated by chi-squared, or Fisher's test as appropriate, comparing percentages in the study population vs Covid19-related cases; MR denotes mortality rates per 100.000 persons-week; CIs denotes confidence intervals for mortality rates and were calculated assuming a Poisson distribution for uncommon events.

**Supplementary Table S7.**
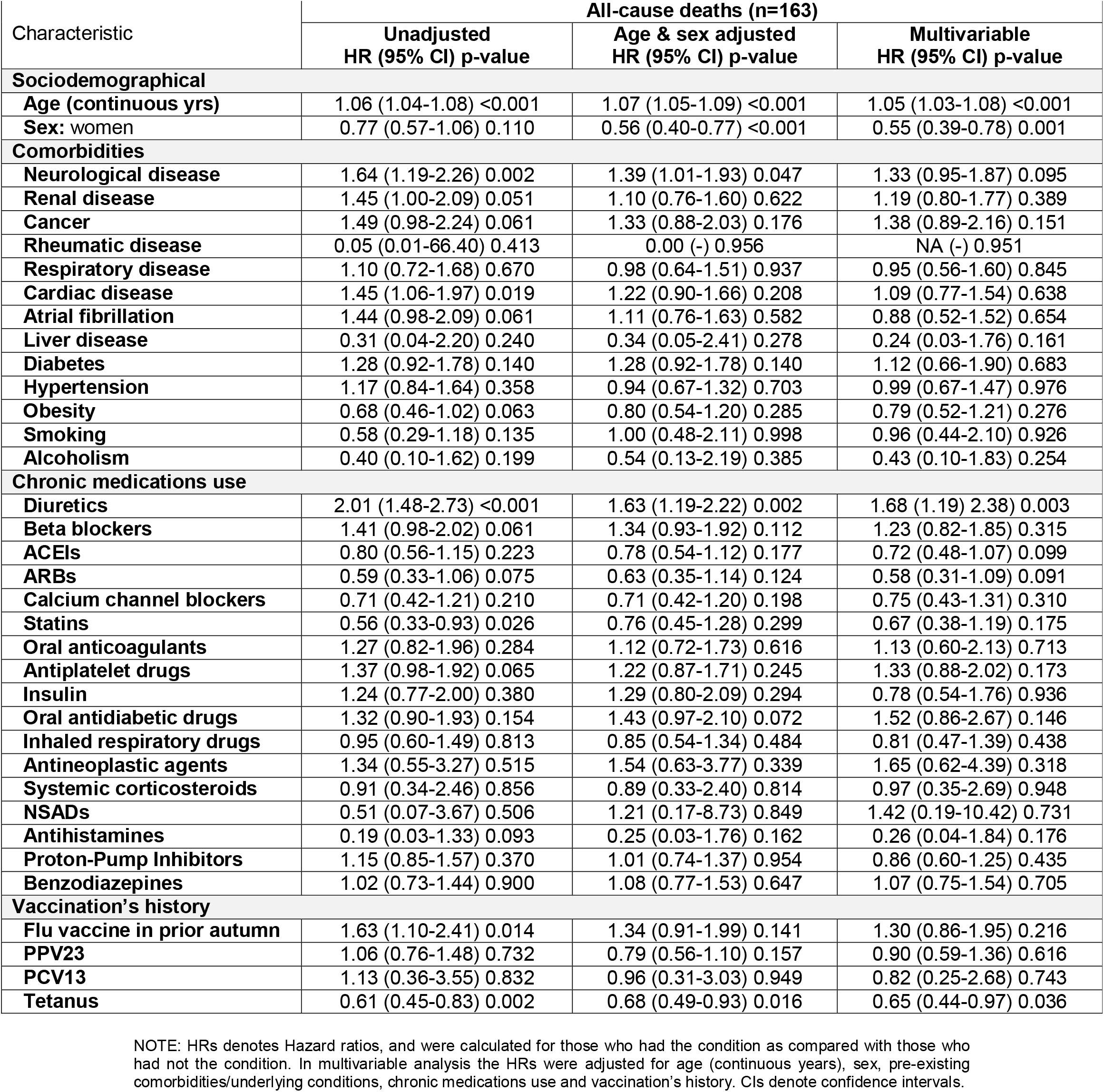
Cox regression analyses assessing unadjusted, age & sex-adjusted and multivariable-adjusted risk of all-cause mortality in nursing-home residents (N=1,414). Tarragona region (Southern Catalonia, Spain) from 01/03/2020 to 30/06/2020.

**Supplementary Table S8.** Cox regression analyses assessing unadjusted, age & sex-adjusted and multivariable-adjusted risk of Covid19-related mortality in nursing-home residents (N=1,414). Tarragona region (Southern Catalonia, Spain) from 01/03/2020 to 30/06/2020.

| Characteristic | COVID-19 deaths (n=61) |  | Multivariable<br>HR (95% CI) p-value |
| --- | --- | --- | --- |
|  | Unadjusted<br>HR (95% CI) p-value | Age & sex adjusted<br>HR (95% CI) p-value |  |
| **Sociodemographical** |  |  |  |
| **Age (continuous yrs)** | 1.05 (1.02-1.08) 0.001 | 1.06 (1.03-1.09) <0.001 | 1.06 (1.03-1.10) <0.001 |
| **Sex: women** | 0.60 (0.36-1.00) 0.049 | 0.45 (0.27-0.76) 0.003 | 0.46 (0.26-0.83) 0.009 |
| **Comorbidities** |  |  |  |
| **Neurological disease** | 1.53 (0.90-2.60) 0.114 | 1.37 (0.80-2.35) 0.248 | 1.19 (0.68-2.08) 0.545 |
| **Renal disease** | 1.01 (0.51-1.99) 0.978 | 0.76 (0.39-1.51) 0.437 | 0.87 (0.42-1.80) 0.714 |
| **Cancer** | 1.47 (0.75-2.90) 0.265 | 1.27 (0.64-2.51) 0.499 | 1.17 (0.56-2.46) 0.677 |
| **Rheumatic disease** | 0.05 (0.01-6912.09) 0.618 | 0.00 (-) 0.973 | 0.00 (-) 0.987 |
| **Respiratory disease** | 0.79 (0.36-1.74) 0.556 | 0.68 (0.31-1.50) 0.342 | 0.67 (0.26-1.71) 0.397 |
| **Cardiac disease** | 1.25 (0.75-2.07) 0.400 | 1.06 (0.64-1.78) 0.814 | 1.18 (0.66-2.11) 0.578 |
| **Atrial fibrillation** | 1.07 (0.54-2.11) 0.847 | 0.83 (0.42-1.64) 0.585 | 1.25 (0.52-2.96) 0.620 |
| **Liver disease** | 0.05 (0.00-169.03) 0.467 | 0.00 (-) 0.962 | 0.00 (-) 0.984 |
| **Diabetes** | 1.39 (0.82-2.36) 0.222 | 1.35 (0.80-2.30) 0.264 | 0.97 (0.38-2.47) 0.950 |
| **Hypertension** | 0.75 (0.45-1.25) 0.272 | 0.63 (0.37-1.05) 0.077 | 0.62 (0.33-1.15) 0.126 |
| **Obesity** | 0.70 (0.36-1.34) 0.277 | 0.81 (0.42-1.55) 0.521 | 0.94 (0.47-1.88) 0.861 |
| **Smoking** | 1.25 (0.54-2.89) 0.609 | 1.86 (0.75-4.62) 0.183 | 2.09 (0.79-5.49) 0.136 |
| **Alcoholism** | 0.54 (0.08-3.92) 0.546 | 0.60 (0.08-4.45) 0.619 | 0.40 (0.05-3.07) 0.376 |
| **Chronic medications use** |  |  |  |
| **Diuretics** | 1.25 (0.74-2.11) 0.397 | 1.05 (0.62-1.78) 0.855 | 1.21 (0.67-2.18) 0.527 |
| **Beta blockers** | 1.00 (0.52-1.92) 0.993 | 0.96 (0.50-1.84) 0.895 | 1.14 (0.56-2.35) 0.718 |
| **ACEIs** | 1.01 (0.58-1.77) 0.974 | 1.00 (0.57-1.74) 0.987 | 0.95 (0.50-1.81) 0.887 |
| **ARBs** | 0.25 (0.06-1.04) 0.056 | 0.28 (0.07-1.13) 0.073 | 0.27 (0.06-1.19) 0.084 |
| **Calcium channel blockers** | 1.05 (0.50-2.22) 0.890 | 1.05 (0.50-2.20) 0.902 | 1.22 (0.54-2.73) 0.633 |
| **Statins** | 0.90 (0.44-1.82) 0.761 | 1.13 (0.55-2.36) 0.736 | 1.13 (0.49-2.61) 0.769 |
| **Oral anticoagulants** | 0.52 (0.19-1.42) 0.202 | 0.44 (0.16-1.22) 0.117 | 0.32 (0.09-1.14) 0.079 |
| **Antiplatelet drugs** | 1.84 (1.04-3.26) 0.036 | 1.25 (0.73-2.16) 0.418 | 1.21 (0.61-2.38) 0.592 |
| **Insulin** | 1.21 (0.55-2.67) 0.631 | 1.22 (0.55-2.68) 0.628 | 0.77 (0.30-1.96) 0.584 |
| **Oral antidiabetic drugs** | 1.44 (0.84-2.47) 0.190 | 1.89 (1.06-3.38) 0.030 | 2.69 (1.02-7.07) 0.045 |
| **Inhaled respiratory drugs** | 0.79 (0.36-1.74) 0.558 | 0.69 (0.31-1.52) 0.357 | 0.85 (0.34-2.11) 0.729 |
| **Antineoplastic agents** | 2.16 (0.68-6.90) 0.193 | 2.41 (0.76-7.71) 0.137 | 3.81 (1.00-14.44) 0.049 |
| **Systemic corticosteroids** | 0.05 (0.00-48.30) 0.389 | 0.00 (-) 0.969 | 0.00 (-) 0.980 |
| **NSADs** | 0.050 (0.00-1882.72) 0.575 | 0.00 (-) 0.972 | 0.00 (-) 0.989 |
| **Antihistamines** | 0.05 (0.00-27.63) 0.348 | 0.00 (-) 0.967 | 0.00 (-) 0.977 |
| **Proton-Pump Inhibitors** | 0.78 (0.46-1.31) 0.345 | 0.68 (0.40-1.14) 0.145 | 0.71 (0.38-1.32) 0.278 |
| **Benzodiazepines** | 1.00 (0.57-1.75) 0.997 | 1.10 (0.62-1.94) 0.744 | 0.88 (0.49-1.60) 0.677 |
| **Vaccination's history** |  |  |  |
| **Flu vaccine in prior autumn** | 2.52 (1.20-5.30) 0.015 | 2.16 (1.02-4.56) 0.043 | 2.34 (1.08-5.07) 0.031 |
| **PPV23** | 0.96 (0.56-1.64) 0.878 | 0.73 (0.42-1.25) 0.245 | 0.74 (0.38-1.47) 0.390 |
| **PCV13** | 0.99 (0.14-7.11) 0.988 | 0.81 (0.11-5.84) 0.832 | 1.07 (0.14-8.27) 0.952 |
| **Tetanus** | 0.61 (0.36-1.01) 0.055 | 0.64 (0.38-1.07) 0.089 | 0.66 (0.34-1.27) 0.210 |

NOTE: HRs denotes Hazard ratios, and were calculated for those who had the condition as compared with those who had not the condition. In multivariable analysis the HRs were adjusted for age (continuous years), sex, pre-existing comorbidities/underlying conditions, chronic medications use and vaccination's history. CIs denote confidence intervals.

